# Opioid prescribing to people on orthopaedic waiting lists during the COVID-19 pandemic in England: a study using OpenSAFELY-TPP

**DOI:** 10.1101/2025.05.06.25326436

**Authors:** Rose Higgins, Rebecca M Smith, Iain Dillingham, Jane Quinlan, Victoria Speed, Helen J Curtis, Chris Wood, Milan Wiedemann, Meghna Jani, Seb C Bacon, Amir Mehrkar, Ben Goldacre, The OpenSAFELY Collaborative, Brian MacKenna, Andrea L Schaffer

**Author notes:** Corresponding author: Dr Andrea Schaffer, Bennett Institute for Applied Data Science, Nuffield Department of Primary Care Health Sciences, University of Oxford.

## Abstract

**Background:** Waiting times for elective procedures increased dramatically during the COVID-19 pandemic. People waiting for orthopaedic procedures often require opioids to manage pain, and longer waiting times may result in a need for more and stronger opioids to treat symptoms.

**Methods:** With NHS England approval, we used routine clinical data from general practice adult patients in OpenSAFELY-TPP linked with the National Minimum Waiting List Dataset. We identified people on elective waiting lists for inpatient trauma or orthopaedic procedures (May 2021-Apr 2022). We quantified changes in weekly opioid prescribing from 6 months pre-waiting list start date to 1 year after the waiting list end date. We also compared long-term opioid prescribing rates in the 3 months before the waiting list start date and months 4-6 after the waiting list end date. We also stratified analyses by time spent on the waiting list (<=18 weeks, 19-52 weeks, >52 weeks).

**Results:** Among 63,850 people on elective trauma or orthopaedic waiting lists (median age = 61 years, 54.6% female), 20.5% waited more than 52 weeks. Weekly opioid prescribing rates per 100 waiting list population were relatively stable over time, with peaks immediately post-treatment, and plateauing again after approximately 3 months. Comparing the 3 months before the waiting list start date to months 4-6 after the waiting list end date, changes in the proportion of people with >=3 opioid prescriptions were -1.6% (95%CI -2.2%, -1.0%) for people on the waiting list <=18 weeks, -1.1% (95%CI -1.7%, -0.5%) among people waiting 19-52 weeks, and -0.5% (95%CI -1.4%, 0.4%) among people waiting >52 weeks.

**Conclusion:** During the COVID-19 pandemic, one in five people waiting for elective orthopaedic procedures waited more than one year. Nearly one in seven were prescribed opioids long-term prior to their referral date, and only small reductions in long-term opioid prescribing was observed post-procedure, regardless of time spent on the waiting list. However, people on waiting lists experienced much longer wait times during the COVID-19 pandemic which also means greater exposure to opioids while awaiting treatment.

## Background

In November 2023, there were 6.4 million patients on elective waiting lists in England. This is a dramatic increase from 4.6 million people in February 2019, due, in part, to healthcare disruptions caused by the pandemic.(1) Some specialties are more greatly affected than others. Wait times for elective surgery including trauma and orthopaedic procedures are well below the care standard of 18 weeks, with almost half of patients waiting for longer.(2) These delays are not experienced equally, with people living in more deprived areas experiencing longer wait times.(3)

Musculoskeletal conditions are a common reason for people to receive elective orthopaedic surgery. Approximately one third of people in the UK have a musculoskeletal condition, such as osteoarthritis, many of whom experience chronic pain.(4) Two of the most common elective inpatient waiting list procedures are knee and hip replacement(5,6) with approximately 200,000 of these procedures performed in the UK in 2018.(7) Longer waiting times may mean patients require stronger analgesia, often opioids, to treat their pain.(8–10) Increased clinical use of opioids may have been exacerbated during the COVID-19 pandemic, when access to non-pharmacological treatments was limited.(11) Major surgery (including orthopaedic surgery) is a known factor associated with long-term opioid use in the UK.(12) In turn, long-term opioid use has been associated with a higher risk of both short- and long-term adverse post-surgical outcomes.(13)

To understand the impact on opioid prescribing of the increase in duration of elective waiting times during the COVID-19 pandemic, we quantified changes in prescribing of opioids to a population with high rates of opioid use (those waiting for trauma or orthopaedic procedures). Specifically, we aimed to: 1) Describe the characteristics of people on elective routine trauma or orthopaedic waiting lists ending in admission during the pandemic; 2) Quantify how patterns of opioid prescribing changed before the waiting list referral date, while on the waiting list, and post-treatment; 3) Establish how these patterns vary by time on the waiting list.

## Methods

### Monthly aggregate waiting list data

Two sources of data relating to waiting lists are available. One contains person-level data on waits (covered in the section below), and the other contains national waiting list data aggregated to specialty and care provider level. Person-level waiting list data were only available between May 2021 and April 2022 (the last month available at the time the analysis was carried out). Therefore, to understand trends pre-COVID-19 pandemic, we used the publicly available monthly aggregate referral-to-treatment (RTT) data for waiting list pathways ending in a hospital admission between January 2019 and June 2022.(14) RTT waiting lists cover waits for consultant-led, elective procedures.(15) A glossary of terms is in Supplementary Box 1.

We restricted the analysis to people waiting for trauma or orthopaedic specialty procedures, as these are commonly performed to treat conditions requiring analgesia.(16,17) Trauma and orthopaedic procedures were defined using the treatment function code ‘110 - Trauma and Orthopaedic Service’, which includes services to treat injuries, congenital and acquired disorders of the bones, joints, and their associated soft tissues, including ligaments, nerves, and muscles.(18,19) It is not possible to distinguish orthopaedic procedures from trauma procedures, so both types of activity were included. However, most trauma procedures are conducted through urgent care pathways and thus were out of scope of the RTT data.

### Person-level waiting list data

#### Data Source

Primary care records managed by the GP software provider, TPP were linked to Office for National Statistics (ONS) death data and the National Minimum Waiting List Dataset (NMWLD) through OpenSAFELY. ONS death data is a longstanding collection whilst the NMWLD was a new data collection rapidly established in response to the pandemic and first approved in March 2021.(20) They were linked, stored and analysed securely within the OpenSAFELY platform: https://opensafely.org/ as part of the NHS England OpenSAFELY COVID-19 service. The data is pseudonymised and includes fields covering coded diagnoses, medications and physiological parameters. No free text data are included. All code is shared openly for review and re-use under MIT open license (https://github.com/opensafely/waiting-list). Detailed pseudonymised patient data are potentially re-identifiable and therefore not shared. A detailed overview of OpenSAFELY is available elsewhere.(21) The NMWLD includes weekly data for people on RTT waiting lists. It is subject to less validation than the official monthly statistics, and therefore the data are less complete.(22)

#### Study population

We identified all patients registered with a GP using TPP electronic health record software who were on a waiting list for an elective (routine, non-urgent) procedure between May 2021 and April 2022. We further restricted the study population to people whose waiting list ended in admission only. Non-admitted pathways include outpatient procedures and people whose waiting time ended for reasons such as declining treatment, entering active monitoring, a decision having been made not to treat, or the patient dies.(15) We could not accurately ascertain the exact reason from the data. Due to this heterogeneity, waits not ending in an admission were excluded.

We excluded people with missing or impossible values of age or sex (<0.01%) as this indicates poor data quality. Extreme waiting times might be spurious outliers; therefore, to reduce their impact on the analysis we also excluded people with values greater than the 99.9th percentile (2.5 years). Finally, we excluded people who were <18 at the start of the waiting list, and with a history of cancer (cancer diagnosis in the five years prior to the waiting list start) as they may be using opioids to treat cancer pain.

For people with more than one eligible pathway, we restricted to the most recently completed pathway. Participants had to be registered with their general practice from 6 months prior to the waiting list start date and through to their waiting list end date. If a patient deregistered from their practice or died during the study period, they were censored. Details of inclusion/exclusion criteria are in **Supplementary Figure 1**.

#### Study measures

The waiting list start date is the date of referral to a consultant-led service and, for patients on an admitted pathway, the waiting list end date is the date of admission, where that admission includes first definitive treatment. We defined key population variables on the waiting list start date: age, sex, Index of Multiple Deprivation (IMD) decile, and ethnicity. For IMD and ethnicity, missing values were included in a separate category. We also identified the presence of the following health conditions: anxiety symptoms; cardiac disease; chronic kidney disease (CKD); chronic respiratory disease; depression symptoms; diabetes; osteoarthritis; rheumatoid arthritis; or severe mental illness (e.g. bipolar disorder, schizophrenia, psychosis). Osteoarthritis and rheumatoid arthritis commonly require analgesia, while mental health problems are also associated with increased opioid use.(23) The other conditions were included as a measure of patients’ general health status.

##### Opioid prescribing

We identified prescribing of all opioids for analgesia in the primary care setting; defined as those falling under the British National Formulary (BNF) Chapters 4.7.2 (Opioid analgesics), in addition to opioids in combination with paracetamol and/or ibuprofen under 4.7.1 and 10.1.1. We excluded opioids not primarily indicated for pain, such as codeine for cough suppression (3.9.1), motility problems (1.4.2), alfentanil / fentanyl for general anaesthesia (15.1.4), and buprenorphine / methadone for opiate substitution therapy (4.10.3).

We also classified opioids based on their strength (weak, moderate, or strong opioids) and duration of action (immediate-release vs modified-release opioids). Weak opioids included codeine, meptazinol, dihydrocodeine; moderate opioids included tramadol and tapentadol; and strong opioids included all other formulations.(24) Immediate-release opioids are preferred for treatment of post-surgical pain, due to the risk of harms associated with modified-release formulations in people receiving surgery.(25,26)

All codelists for medicines and comorbidities in this study are available in our Github repository: https://github.com/opensafely/waiting-list.

##### Other analgesic prescribing

We identified the prescribing of other medicines to treat pain in the 3 months prior to waiting list start date, specifically: antidepressants including those indicated for treatment of neuropathic pain (amitriptyline, duloxetine)(27); gabapentinoids; and non-steroidal antiinflammatory drugs (NSAIDs).

#### Analysis

We took two approaches to understanding changes in opioid prescribing to people on a waiting list: first, we performed a before and after comparison of opioid prescribing in the period immediately prior to the waiting list referral date, and after the waiting list end date (i.e. admission for treatment). Second, we looked at changes in prescribing patterns by week throughout the study period. We did this for overall opioid prescribing, and for different types (weak, moderate or strong opioids; immediate-release or modified-release opioids). To prevent disclosure, all counts are rounded to the nearest 5.

##### I. Before and after analysis

We calculated the number of people with ≥1 and ≥3 opioid prescriptions in the 3 months before the waiting list referral date and in months 4-6 post-waiting list end date. We looked at prescribing of ≥3 opioid prescriptions in 3 months as it has previously been used to define long-term opioid use in UK routine data.(28)

For the post-treatment time period, we excluded the first 3 months after the waiting list end date and focussed on months 4-6. We did this to exclude the time period immediately post-treatment where people were using opioids temporarily to treat pain from their procedure. Furthermore, Royal College of Anaesthetists guidelines state that opioid use beyond 3 months post-surgery warrants further assessment.(25) This is visualised in **Supplementary Figure 2**. To calculate rates of prescribing in the post-waiting list period, we only included people who had full follow-up (i.e. who were still alive and registered with the same practice).

##### II. Changes over time

We quantified the opioid prescribing rate (per 100 people) for each week from 26 weeks prior to the waiting list start date, throughout time on the waiting list, and up to 52 weeks after the waiting list end date. For visualisation in figures, time on the waiting list was truncated at 52 weeks.

In each week, the numerator was the number of people prescribed an opioid in that week, and the denominator was all people still on the waiting list. For instance, for calculating the opioid prescribing rate for week 18 on the waiting list, only people with waits ≥18 weeks were included.

To understand how prescribing patterns varied by time on the waiting list, we also stratified by waiting list duration (≤18 weeks, 19-52 weeks, >52 weeks). These categories were chosen as ≤18 weeks is considered the waiting list standard for non-urgent procedures, and NHS England policy states that no one should wait for more than 52 weeks from referral to first treatment.(29) To better visualise the non-linear changes in the prescribing rate over time, we smoothed the data using loess regression.

#### Subgroup analyses

As we could not ascertain the specific reason that patients were on the waiting list, we performed several subgroup analyses. First, we restricted our analysis to people with a recorded diagnosis of osteoarthritis in primary care in the past 5 years. Second, we identified people who had received an orthopaedic hip procedure, or knee procedure, by linking to Hospital Episode Statistics Admitted Patient Care (HES APC) data and identified using Healthcare Resource Groups (HRG) codes (**Supplementary Table 1**).

#### Ethics approval

This study was approved by the Health Research Authority (Research Ethics Committee reference 20/LO/0651) and by the London School of Hygiene and Tropical Medicine Ethics Board (reference 21863).

## Results

### Monthly aggregate waiting list data

In the year prior to the COVID-19 pandemic (March 2019 to February 2020), there were 587,977 people admitted for elective trauma or orthopaedic treatment, with a median of 49,253 admissions per month (**Figure 1A**). Overall, during this year the proportion of people who waited more than 52 weeks was 0.5% (n=3046) (**Figure 1B**).

**Figure 1.**
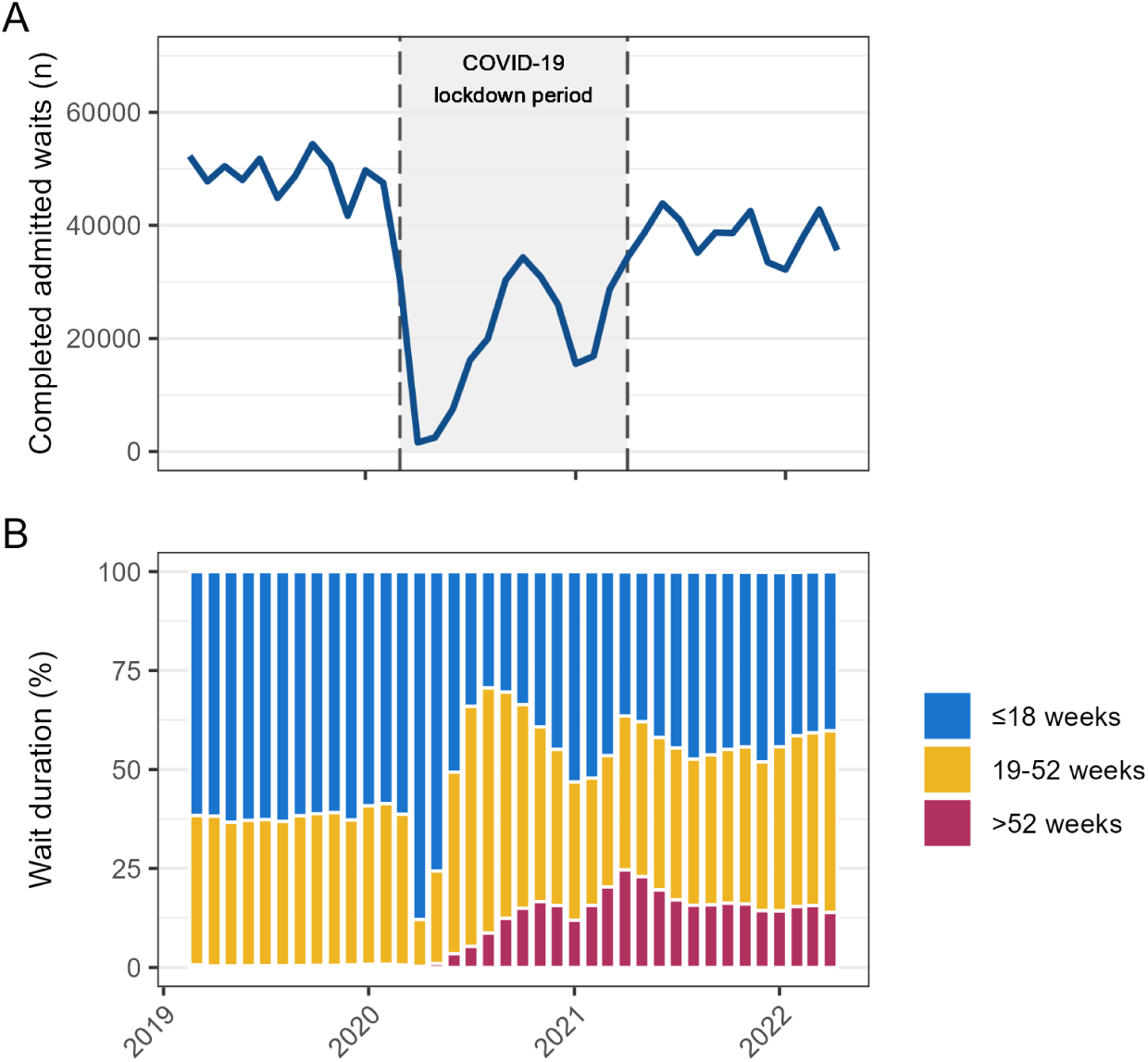
Number of completed waits for elective trauma and orthopaedic admitted procedures based on month of waiting list end date (A) and time on waiting list for these procedures (B), Jan 2019 to Apr 2022.

Following decreases in admissions of patients on the waiting list during the COVID-19 lockdowns (Mar 2020 to Apr 2021), from May 2021 to Apr 2022 there were 460,830 elective trauma or orthopaedic admissions with a median of 38,688 per month. A total of 16.5% (n=76,075) had waited >52 weeks before being admitted for treatment, with 56.7% (n=260,762) waiting >18 weeks.

### Person-level waiting list data

In the person-level waiting list data, there were nearly 3 million people who were on an RTT waiting list that ended between May 2021 to April 2022. Of these, 235,540 were waiting for trauma or orthopaedic specialty treatment, of which 85.6% were elective (not urgent). After restricting to people waiting for elective inpatient treatment and applying our exclusion criteria, we identified 63,850 people in our study cohort (**Table 1, Supplementary Figure 1**). Of these, 6,150 (9.6%) were on more than one eligible waiting list pathway. The waiting list referral and end dates are in **Supplementary Figure 3**. The median age was 61 years (interquartile range [IQR], 49-72) and 54.6% were female. The median time spent on the waiting list was 26 weeks (IQR 12-47); 24,390 (38.2%) waited 18 weeks or less, 26,375 (41.3%) 19-52 weeks, and 13,090 (20.5%) waited more than 52 weeks.

**Table 1.** Characteristics of people waiting for elective inpatient trauma or orthopaedic procedures between 1 May 2021 and 30 Apr 2022, overall and stratified by opioid prescribing in the 3 months period to the waiting list start date.

|  | Full cohort | 3 months prior to waiting list referral date |  |  |
| --- | --- | --- | --- | --- |
|  |  | No opioid prescription | ≥1 prescription | ≥3 prescriptions |
|  | n (%) | n (%) | n (%) | n (%) |
| **Total** | 63850 (100.0) | 43,275 (100.0) | 20,575 (100.0) | 9,890 (100.0) |
| **Age on waiting list start date** |  |  |  |  |
| 18-39 | 8915 (14.0) | 7545 (17.4) | 1,380 (6.7) | 510 (5.2) |
| 40-49 | 7165 (11.2) | 5180 (12.0) | 1,985 (9.6) | 940 (9.5) |
| 50-59 | 13530 (21.2) | 9105 (21.0) | 4,450 (21.6) | 2,185 (22.1) |
| 60-69 | 15085 (23.7) | 9495 (21.9) | 5,600 (27.2) | 2,815 (28.5) |
| 70-79 | 14180 (22.2) | 8985 (20.8) | 5,215 (25.3) | 2,495 (25.2) |
| 80+ | 4905 (7.7) | 2965 (6.9) | 1,940 (9.4) | 940 (9.5) |
| **Sex** |  |  |  |  |
| Female | 34820 (54.6) | 22295 (51.5) | 12,565 (61.1) | 6,195 (62.6) |
| Male | 28960 (45.4) | 20980 (48.5) | 8,005 (38.9) | 3,695 (37.4) |
| **IMD decile** |  |  |  |  |
| 1 (most deprived) | 5310 (8.3) | 3105 (7.2) | 2,205 (10.7) | 1,210 (12.2) |
| 2 | 5790 (9.1) | 3550 (8.2) | 2,245 (10.9) | 1,200 (12.1) |
| 3 | 5670 (8.9) | 3590 (8.3) | 2,080 (10.1) | 1,075 (10.9) |
| 4 | 6260 (9.8) | 4140 (9.6) | 2,125 (10.3) | 1,040 (10.5) |
| 5 | 6730 (10.6) | 4565 (10.5) | 2,170 (10.5) | 1,035 (10.5) |
| 6 | 7500 (11.8) | 5210 (12) | 2,300 (11.2) | 1,050 (10.6) |
| 7 | 6940 (10.9) | 4870 (11.3) | 2,080 (10.1) | 970 (9.8) |
| 8 | 6495 (10.2) | 4630 (10.7) | 1,875 (9.1) | 825 (8.3) |
| 9 | 6550 (10.3) | 4795 (11.1) | 1,760 (8.6) | 760 (7.7) |
| 10 (least deprived) | 5345 (8.4) | 3985 (9.2) | 1,365 (6.6) | 545 (5.5) |
| Missing | 1195 (1.9) | 830 (1.9) | 365 (1.8) | 180 (1.8) |
| **Ethnicity** |  |  |  |  |
| White | 46990 (73.7) | 31580 (73) | 15,470 (75.2) | 7,560 (76.4) |
| Black | 1600 (2.5) | 1170 (2.7) | 435 (2.1) | 160 (1.6) |
| South Asian | 615 (1.0) | 435 (1) | 180 (0.9) | 60 (0.6) |
| Mixed | 345 (0.5) | 255 (0.6) | 90 (0.4) | 30 (0.3) |
| Other | 345 (0.5) | 275 (0.6) | 70 (0.3) | 25 (0.3) |
| Unknown | 13875 (21.8) | 9560 (22.1) | 4,325 (21) | 2,060 (20.8) |
| **Conditions recorded in primary care in past 5 years** |  |  |  |  |
| Anxiety (symptoms or diagnosis) | 7730 (12.1) | 5015 (11.6) | 2730 (13.3) | 1385 (14.0) |
| Cardiac disease | 6880 (10.8) | 4010 (9.3) | 2880 (14) | 1545 (15.6) |
| Chronic kidney disease | 2255 (3.5) | 1350 (3.1) | 910 (4.4) | 470 (4.8) |
| Chronic respiratory disease | 2780 (4.4) | 1405 (3.2) | 1385 (6.7) | 815 (8.2) |
| Depression (symptoms or diagnosis) | 8410 (13.2) | 5060 (11.7) | 3370 (16.4) | 1820 (18.4) |
| Diabetes | 5975 (9.4) | 3350 (7.7) | 2625 (12.8) | 1410 (14.3) |
| Osteoarthritis | 24225 (38.0) | 13910 (32.1) | 10345 (50.3) | 5160 (52.2) |
| Rheumatoid arthritis | 1255 (2.0) | 630 (1.5) | 630 (3.1) | 365 (3.7) |
| **Prescribing 3 months prior to waiting list referral date** |  |  |  |  |
| Antidepressants |  |  |  |  |
| ≥1 prescription | 17180 (26.9) | 8385 (19.4) | 8795 (42.7) | 5120 (51.8) |
| ≥3 prescriptions | 9760 (15.3) | 4195 (9.7) | 5565 (27.0) | 3850 (38.9) |
| Amitriptyline/duloxetine |  |  |  |  |
| ≥1 prescription | 7650 (12.0) | 3100 (7.2) | 4550 (22.1) | 2760 (27.9) |
| ≥3 prescriptions | 3720 (5.8) | 1255 (2.9) | 2465 (12.0) | 1860 (18.8) |
| Gabapentinoids |  |  |  |  |
| ≥1 prescription | 6335 (9.9) | 2250 (5.2) | 4085 (19.9) | 2805 (28.4) |
| ≥3 prescriptions | 4020 (6.3) | 1230 (2.8) | 2790 (13.6) | 2215 (22.4) |
| NSAIDs |  |  |  |  |
| ≥1 prescription | 10275 (16.1) | 4975 (11.5) | 5300 (25.8) | 2605 (26.3) |
| ≥3 prescriptions | 3205 (5.0) | 1185 (2.7) | 2020 (9.8) | 1425 (14.4) |
| **Died in 6 months following waiting list end date** | 280 (0.4) | 135 (0.3) | 180 (0.9) | 85 (0.9) |

#### I. Before and after analysis

Approximately one third (n = 20,575, 32.2%) were prescribed an opioid in the 3 months prior to the waiting list referral date (**Table 1**), and 15.5% (n = 9,890) had three or more opioid prescriptions. Immediate-release opioids were more commonly prescribed (29.7% of population) compared with modified-release (5.2%) (**Table 2**). 23.7% were prescribed a weak opioid, compared with 6.6% a moderate opioid and 5.9% a strong opioid.

**Table 2.** Opioid prescribing before and after time on waiting list among people on elective inpatient trauma or orthopaedic waiting lists, by time on waiting list (<=18 weeks, 19-52 weeks, >52 weeks) (n= 63,850). “Pre-waiting list” is 3 months prior to referral date, and “Post-waiting list” is months 4-6 after waiting list end date.

|  | ≥1 opioid prescription |  |  | ≥3 opioid prescriptions |  |  |
| --- | --- | --- | --- | --- | --- | --- |
|  | Pre-waiting list | Post-waiting list<sup>^</sup> | Difference | Pre-waiting list | Post-waiting list<sup>^</sup> | Difference |
|  | n (%) | n (%) | % (95% CI) | n (%) | n (%) | % (95% CI) |
| **Full cohort** |  |  |  |  |  |  |
| Any opioid | 20,575 (32.2) | 16,220 (26.9) | -5.4 (-5.8, -4.8) | 9890 (15.5) | 8650 (14.3) | -1.2 (-1.6, -0.8) |
| Immediate-release opioid | 18,970 (29.7) | 14,650 (24.3) | -5.4 (-5.9, -4.9) | 8135 (12.7) | 7010 (11.6) | -1.1 (-2.1, -0.2) |
| Modified-release opioid | 3,330 (5.2) | 3,055 (5.1) | -0.2 (-0.4, 0.1) | 2300 (3.6) | 2245 (3.7) | 0.1 (-0.5, 0.7) |
| Weak opioid | 15,150 (23.7) | 11,195 (18.5) | -5.2 (-5.8, -4.7) | 5655 (8.9) | 4865 (8.1) | -0.8 (-2.4, 0.8) |
| Moderate opioid | 4,210 (6.6) | 3,370 (5.6) | -1.0 (-1.3, -0.7) | 2000 (3.1) | 1800 (3.0) | -0.2 (-0.8, 0.5) |
| Strong opioid | 3,790 (5.9) | 3,480 (5.8) | -0.2 (-0.4, 0.1) | 4385 (6.9) | 4095 (6.8) | -0.08 (-1.7, 1.5) |
| **Time on waiting list: <=18 weeks** |  |  |  |  |  |  |
| Any opioid | 7660 (31.4) | 5925 (25.8) | -5.6 (-6.4, -4.8) | 3785 (15.5) | 3195 (13.9) | -1.6 (-2.2, -1) |
| Immediate-release opioid | 7035 (28.8) | 5340 (23.2) | -5.6 (-6.4, -4.8) | 3110 (12.8) | 2565 (11.2) | -1.6 (-2.2, -1) |
| Modified-release opioid | 1315 (5.4) | 1160 (5) | -0.4 (-0.8, 0) | 920 (3.8) | 865 (3.8) | 0.0 (-0.3, 0.3) |
| Weak opioid | 5485 (22.5) | 4030 (17.5) | -5 (-5.7, -4.3) | 2085 (8.6) | 1760 (7.7) | -0.9 (-1.4, -0.4) |
| Moderate opioid | 1605 (6.6) | 1280 (5.6) | -1 (-1.4, -0.6) | 790 (3.2) | 685 (3) | -0.2 (-0.5, 0.1) |
| Strong opioid | 1590 (6.5) | 1335 (5.8) | -0.7 (-1.1, -0.3) | 1020 (4.2) | 930 (4) | -0.2 (-0.6, 0.2) |
| **Time on waiting list: 19-52 weeks** |  |  |  |  |  |  |
| Any opioid | 8540 (32.4) | 6695 (26.8) | -5.6 (-6.4, -4.8) | 3985 (15.1) | 3505 (14) | -1.1 (-1.7, -0.5) |
| Immediate-release opioid | 7935 (30.1) | 6070 (24.3) | -5.8 (-6.6, -5) | 3315 (12.6) | 2875 (11.5) | -1.1 (-1.7, -0.5) |
| Modified-release opioid | 1280 (4.9) | 1190 (4.8) | -0.1 (-0.5, 0.3) | 875 (3.3) | 865 (3.5) | 0.2 (-0.1, 0.5) |
| Weak opioid | 6445 (24.4) | 4705 (18.8) | -5.6 (-6.3, -4.9) | 2380 (9) | 2025 (8.1) | -0.9 (-1.4, -0.4) |
| Moderate opioid | 1680 (6.4) | 1325 (5.3) | -1.1 (-1.5, -0.7) | 765 (2.9) | 700 (2.8) | -0.1 (-0.4, 0.2) |
| Strong opioid | 1425 (5.4) | 1360 (5.4) | 0 (-0.4, 0.4) | 930 (3.5) | 920 (3.7) | 0.2 (-0.1, 0.5) |
| **Time on waiting list: >52 weeks** |  |  |  |  |  |  |
| Any opioid | 4380 (33.5) | 3610 (29.1) | -4.4 (-5.5, -3.3) | 2120 (16.2) | 1945 (15.7) | -0.5 (-1.4, 0.4) |
| Immediate-release opioid | 4000 (30.6) | 3240 (26.2) | -4.4 (-5.5, -3.3) | 1710 (13.1) | 1570 (12.7) | -0.4 (-1.2, 0.4) |
| Modified-release opioid | 735 (5.6) | 705 (5.7) | 0.1 (-0.5, 0.7) | 505 (3.9) | 515 (4.2) | 0.3 (-0.2, 0.8) |
| Weak opioid | 3220 (24.6) | 2460 (19.9) | -4.7 (-5.7, -3.7) | 1190 (9.1) | 1080 (8.7) | -0.4 (-1.1, 0.3) |
| Moderate opioid | 925 (7.1) | 765 (6.2) | -0.9 (-1.5, -0.3) | 450 (3.4) | 410 (3.3) | -0.1 (-0.5, 0.3) |
| Strong opioid | 775 (5.9) | 790 (6.4) | 0.5 (-0.1, 1.1) | 510 (3.9) | 545 (4.4) | 0.5 (0, 1) |

\*Excluding people lost to follow-up

Compared with people not prescribed opioids, people with ≥3 opioid prescriptions were more likely to be older, female, living in more deprived areas, and have a diagnosis of osteoarthritis (52.2% vs 32.1%). They were also much more likely to be prescribed antidepressants and other analgesic medicines (gabapentinoids, NSAIDs).

Comparing months 4-6 after the end of the waiting list to the 3 months prior to the waiting list referral date, there were slight reductions in the proportion of people prescribed immediate-release opioids (-5.4%, 95%CI -5.8, -4.8) but prescribing of modified-release opioids did not change (-0.2%, 95%CI -0.4, 0.1). Similarly, fewer people were prescribed weak opioids (-5.2%, 95%CI -5.8, -4.7) but only slight changes were observed for moderate and strong opioids. There was substantial variation in opioid prescribing by age, sex, and IMD decile (**Table 1**).

The proportion of people with ≥3 opioid prescriptions changed from 15.5% in the 3 months pre-waiting list to 14.3% post-waiting list (-1.2%, 95%CI -1.6, -0.8) (**Table 2**). When stratified by time on the waiting list, similar patterns were observed. There was slight variation in the opioid prescribing rate pre-waiting list, ranging from 31.4% and 15.5% for ≥1 and ≥3 opioid prescriptions for people waiting ≤18 weeks, to 33.5% and 16.2% for people waiting >52 weeks. In months 4-6 post-treatment, these values ranged from 25.8% and 13.9% to 29.1% and 15.7% (**Table 2**). The reduction in proportion of people with ≥3 opioid prescriptions ranged from -0.5% for people waiting >52 weeks, to -1.6% for people waiting ≤18 weeks.

#### II. Changes in weekly opioid prescribing over time

In the 6 months pre-waiting list, the median number of prescriptions per week was 6.6 per 100 people at 26 weeks prior to the waiting list referral date, increasing to 8.7 the week immediately prior to the referral date (**Figure 2A**). During the period on the waiting list, the median weekly prescribing rate was 7.7 per 100 people (IQR 7.5, 7.9).

**Figure 2.**
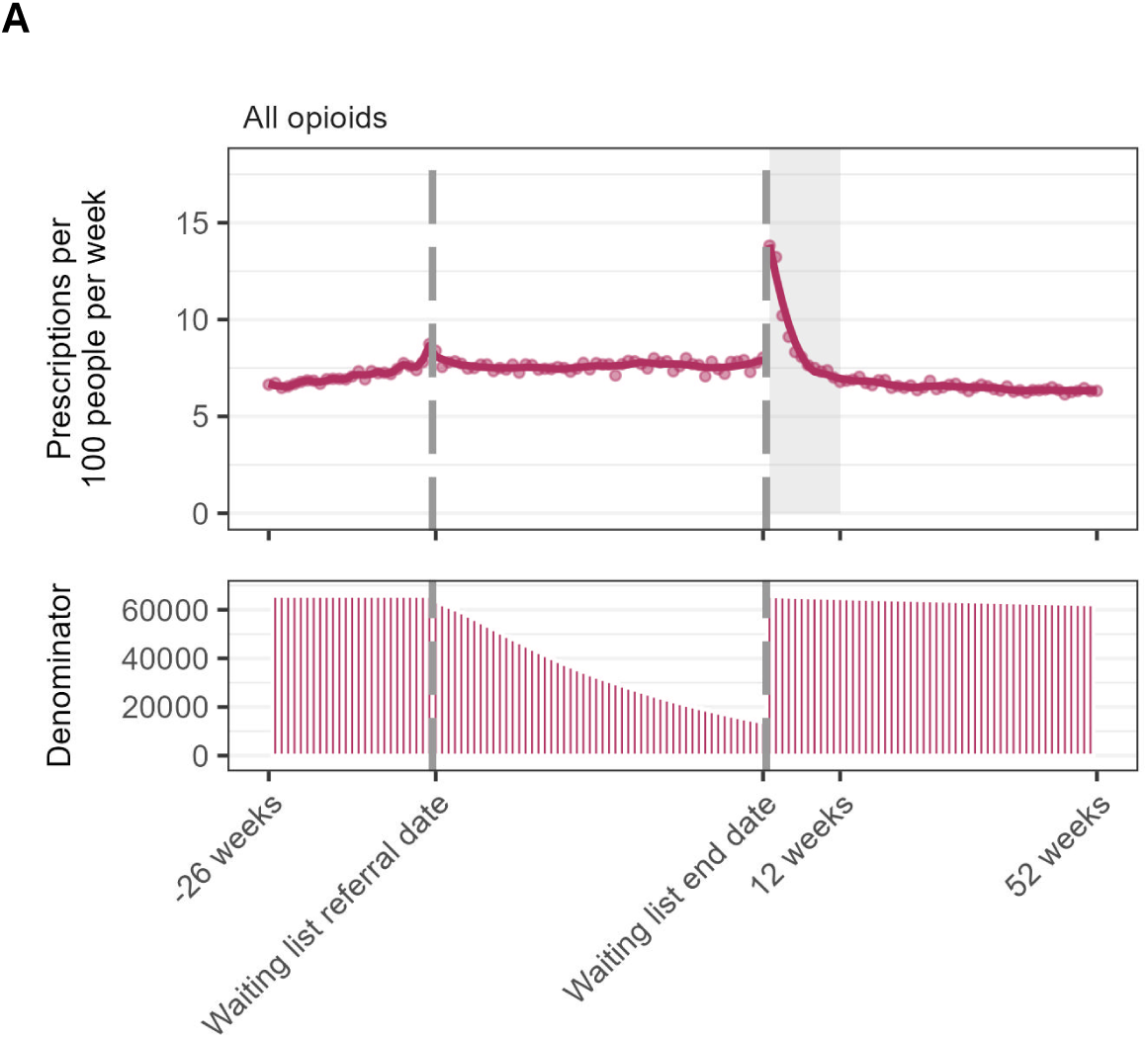
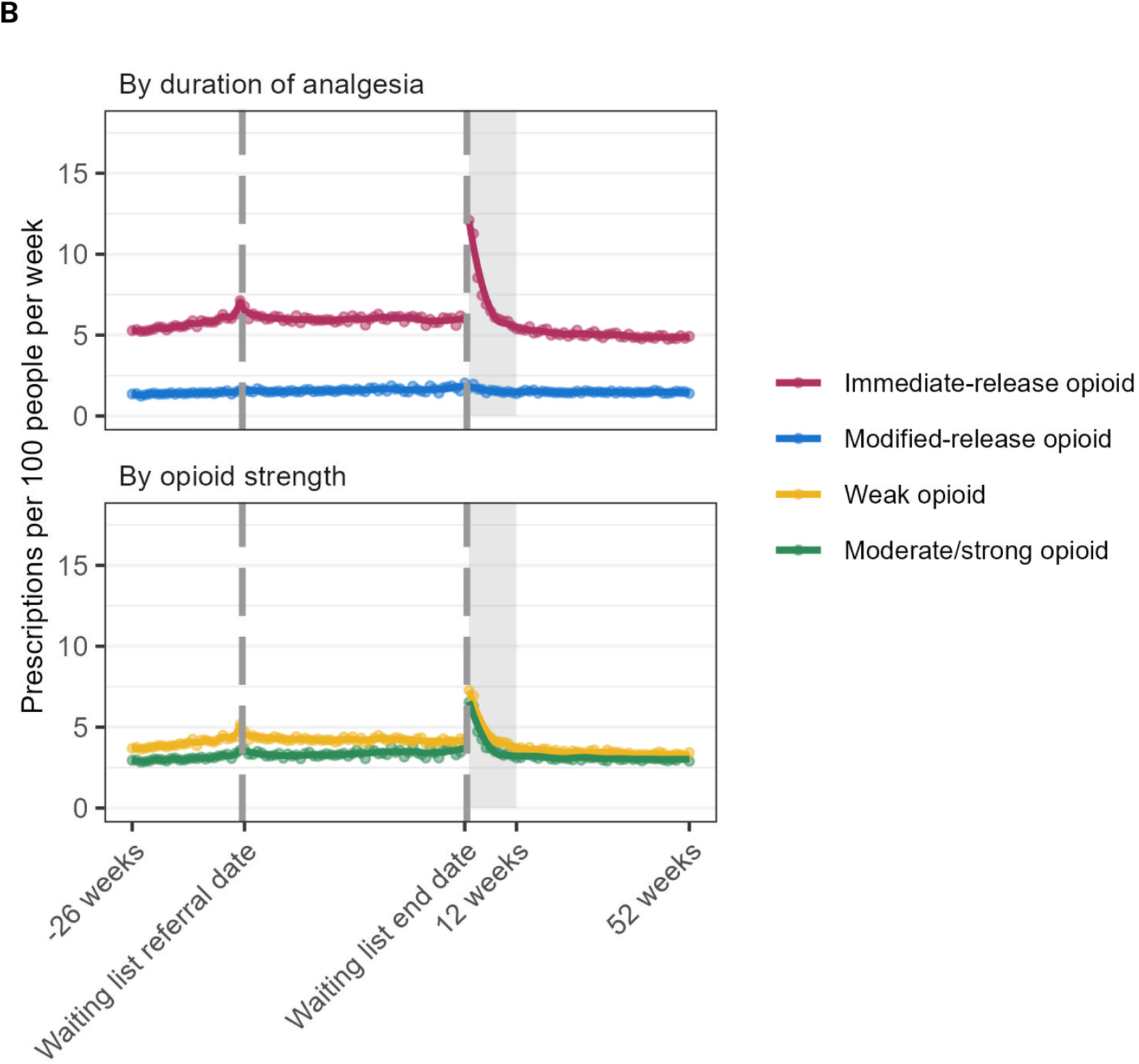
Number of opioid prescriptions per week per 100 people in the 26 weeks prior to waiting list referral date, during the waiting list, and up to 52 weeks after waiting list end date (date of treatment) overall (A) and by opioid type (B). Dots are observed values, solid lines are values predicted from loess regression model. Shaded area is 3 months post-treatment. Legend: During the waiting list period, the denominator includes everyone who was still on the waiting list at the end of each week. During the waiting list period and post-treatment periods, the denominator excludes people who died or who deregistered from their general practice.

There was a large increase in prescribing immediately post-treatment (13.8 per 100 people), which stabilised after approximately 3 months. From 3 months through one year post-waiting list, the median number of prescriptions was 6.5 per 100 people per week (**Figure 2A**). When stratified by opioid type, the greatest changes were observed for immediate-release opioids (**Figure 2B**).

Stratified by time on the waiting list, in the 6 months pre-waiting list the median prescribing rates ranged from 6.8 to 7.2 per 100 people per week. In the 4-6 months post-waiting list end date, the median weekly prescribing rates ranged from 6.4 to 6.9 per 100 people (**Figure 3**). We observed similar patterns by overall total time on the waiting list. In all groups, prescribing stabilised after approximately 3 months.

**Figure 3.**
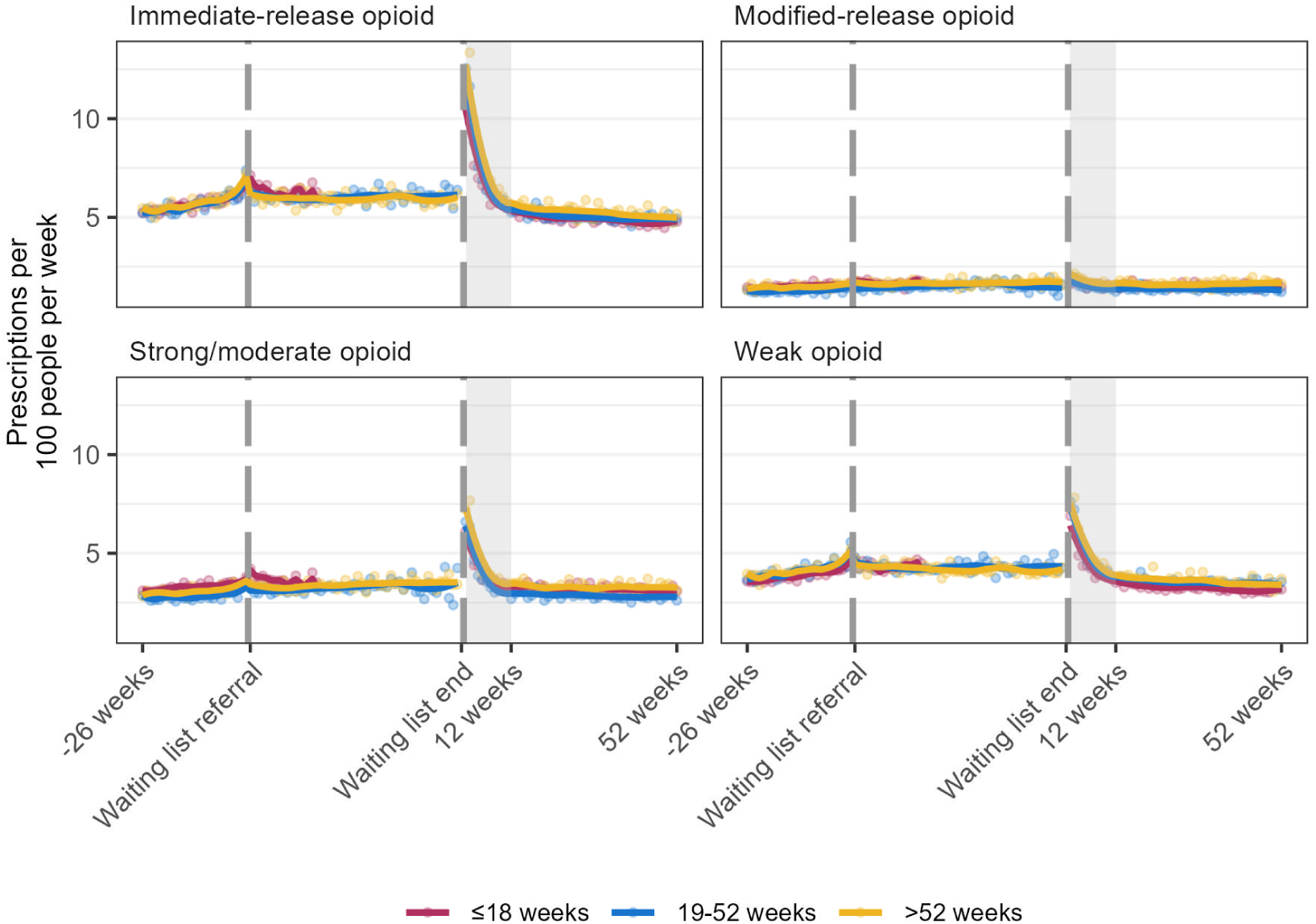
Number of opioid prescriptions per week per 100 people in the 26 weeks prior to waiting list referral date, during the waiting list, and 52 weeks after waiting list end date (date of treatment) stratified by time on waiting list and by opioid type. Dots are observed values, solid lines are values predicted from loess regression model. Shaded area is 3 months post-admission. Legend: During the waiting list period, the denominator includes everyone who was still on the waiting list at the end of each week. During the waiting list period and post-treatment periods, the denominator excludes people who died or who deregistered from their general practice.

### Subgroup analyses

In our sensitivity analyses, there were 24,255 with an osteoarthritis diagnosis, 10,160 who had a hip procedure, and 14,975 with a knee procedure. People who had an inpatient knee procedure were younger, and more likely to be male (**Supplementary Table 2**). Before the waiting list referral date, people with osteoarthritis and a hip procedure had the highest proportion of people with ≥3 opioid prescriptions (21.3% and 21.5%). People with a hip procedure experienced the greatest reduction in opioid prescribing comparing before and after the waiting list: from 44.5% to 30.2% for >=1 prescriptions, and from 21.5% to 15.8% for ≥3 prescriptions (**Supplementary Table 3**). These patterns were mirrored in the plots showing changes in weekly prescribing over time (**Supplementary Figures 4-6**).

## Discussion

### Summary

In the period prior to the COVID-19 pandemic, waits of over 52 weeks for elective trauma or orthopaedic procedures were incredibly rare at 0.5%. However, during our study period during the COVID-19 pandemic, this previously small population increased 25-fold. In addition, more than half of all waits exceeded the 18-week NHS treatment target. In our study population, approximately one third were prescribed an opioid in the three months leading up to their referral date, and nearly one in seven had 3 or more prescriptions, sometimes considered long-term use,(28) with minimal changes post-treatment. While similar patterns of opioid prescribing post-treatment were observed regardless of time on the waiting list, people on waiting lists experienced much longer wait times during the COVID-19 pandemic which also means greater exposure to opioids while awaiting treatment.(30)

### Findings in context

Other studies of changes in opioid prescribing during the pandemic saw no sustained increases in opioid prescribing, either in the general population or in people with musculoskeletal diseases.(31,32) However, a Scottish study of people receiving hip and knee arthroplasty found that opioid use post-surgery was higher during the COVID-19 pandemic than in historic controls.(8) While we observed only slight reductions in opioid use post-procedure, these were mostly among people with no evidence of long-term use (<3 prescriptions) or prescribed weak opioids.

Many orthopaedic procedures (e.g. knee and hip replacements) are often performed with the goal of reducing pain, which is often the indication for the procedure. Thus it is interesting that we did not see greater reductions in prescribing post-procedure. One potential explanation is that osteoarthritis may affect more than the primary joint. Prolonged preoperative opioid is also the greatest risk factor for long-term post-operative use.(33) Studies from Finland(9) and Norway(10) similarly found only modest reductions in opioid prescribing among people with knee and hip arthroplasty, and also observed greater reduction in people with a hip procedure as compared with knee procedure.(9)

It should be noted that there is no standard way of defining long-term use in routine data, with definitions varying depending on study purpose, data availability, and jurisdiction.(34) We used three prescriptions in 3 months, which has been used previously in UK primary care data.(28) However, other UK studies in primary care have defined it variably, including as 3 prescriptions in 1 year.(35)

### Strengths and limitations

We had data for a representative sample of approximately 40% of the English population(36). We could not compare our findings with people on waiting lists pre-COVID-19 as the NMWLD is a new dataset established in response to the COVID-19 pandemic. However we did compare opioid prescribing rates stratified by individuals’ time on the waiting list to better understand how longer waits impacted on opioid prescribing. Waits beyond one year for trauma or orthopaedic procedures were extremely rare prior to the pandemic,(14) but very common in our pandemic cohort.

Our analysis is descriptive and people with shorter waits may be systematically different from those waiting longer, in ways that we could not ascertain (e.g. type of procedure, health status) and that may be associated with propensity for opioid use. Further, the health condition that was being treated and treatment received was not well recorded in the waiting list data. We performed sensitivity analyses among people with evidence of hip and/or knee procedures, but we had to rely on linkage with hospital admissions data which can have a degree of error(37–39).

We were interested in the impact of waiting list length on opioid prescribing. As we did not have information on indication for prescribing, to understand how waiting list length impacted opioid prescribing among people waiting for orthopaedic procedures it was therefore necessary to exclude patients with other conditions which may be driving opioid prescribing. One potential reason for the limited reduction in opioid prescribing post-procedure is that some patients were using opioids for other conditions. While we excluded people with a history of cancer, it is possible other chronic pain conditions may have driven some opioid prescribing and would have been minimally impacted by treatment.

The person-level waiting list data is restricted to primary care prescribing only. However the lack of secondary care prescribing data is not relevant, as long-term opioid prescribing would occur in primary care settings.

### Policy implications

Since the increase in waiting times starting during the COVID-19 pandemic, reducing elective waiting lists has been a government priority.(40) In mid-2024, the UK government pledged to clear the NHS waiting list backlog in 5 years, by increasing the number of appointments.(41) There are many negative impacts of long wait times, which include unnecessary exposure to opioid analgesia. The risks of opioid prescribing are well-established, and use of these medicines should be minimised as much as possible. In 2023, NHS England issued instructions to improve opioid use, highlighting the need for better use of data to prevent and reduce opioid harm.(42)

Waiting lists are a key national priority, however we are unaware of any other studies using the new NMWLD patient-level information to inform interventions and policy-making.(43) We have demonstrated how use of person-level waiting list data, linked to general practice data within OpenSAFELY can be used to better understand opioid prescribing in a high risk population in line with key recommendations. We are developing tools to facilitate near real-time audit and feedback in the context of rapidly evolving pressures on the health service. These can include any measures on opioids which would support NHS England’s ambition on safe opioid use and interventions to support reductions in increased waiting lists driven by the COVID-19 pandemic.

### Future research

There may be heterogeneity in our findings that we have not identified. More research into which patients are most likely to continue with long-term opioid use or experience other opioid-related harms after inpatient procedures, and how this changed during the pandemic, is needed. For instance, an incidental finding was very high rates of prescribing of other analgesic medicines, such as gabapentinoids and antidepressants despite a lack of evidence for use in people with osteoarthritis(44), and increased risk of harms associated with co-prescribing.(45) The impact of longer wait times resulting from COVID-19-related disruptions is likely to extend beyond greater analgesic prescribing, to both functional and psychological impacts.(46,47) Longer waits are also associated with poorer outcomes for hip or knee arthroplasty or joint replacements.(46) More broadly, the NMWLD linked to general practice records offers a new opportunity to understand how the NHS delivers care.

### Conclusion

At a population level, while there may be multiple factors influencing the rapid increase in waiting list times during COVID and the population on a waiting list, this population did not exist to the same extent before the COVID-19 pandemic. Understanding the resource and clinical needs of this new, large cohort is needed to develop robust policy around reducing the waitlist and preventing future recurrence.

The majority of people waiting for routine orthopaedic procedures waited more than the standard 18 weeks, and one in five waited more than one year, which was practically non-existent pre-pandemic. We saw no evidence that longer times spent on the waiting list, as experienced during the COVID-19 pandemic, impacted on prescribing patterns post-treatment. However, people on waiting lists experienced much longer wait times during the COVID-19 pandemic which also means greater exposure to opioids while awaiting treatment.

## Data Availability

Access to the underlying identifiable and potentially re-identifiable pseudonymised electronic health record data is tightly governed by various legislative and regulatory frameworks, and restricted by best practice. The data in the NHS England OpenSAFELY COVID-19 service is drawn from General Practice data across England where EMIS and TPP are the data processors. EMIS and TPP developers initiate an automated process to create pseudonymised records in the core OpenSAFELY database, which are copies of key structured data tables in the identifiable records. These pseudonymised records are linked onto key external data resources that have also been pseudonymised via SHA-512 one-way hashing of NHS numbers using a shared salt. University of Oxford, Bennett Institute for Applied Data Science developers and PIs, who hold contracts with NHS England, have access to the OpenSAFELY pseudonymised data tables to develop the OpenSAFELY tools. These tools in turn enable researchers with OpenSAFELY data access agreements to write and execute code for data management and data analysis without direct access to the underlying raw pseudonymised patient data, and to review the outputs of this code. All code for the full data management pipeline - from raw data to completed results for this analysis - and for the OpenSAFELY platform as a whole is available for review at github.com/OpenSAFELY.

## Acknowledgements

We are very grateful for all the support received from the TPP Technical Operations team throughout this work, and for generous assistance from the information governance and database teams at NHS England and the NHS England Transformation Directorate.

Members of the The OpenSAFELY Collaborative include: Sebastian CJ Bacon, Lucy Bridges, Benjamin FC Butler-Cole, Simon Davy, Iain Dillingham, David Evans, Louis Fisher, Amelia Green, Ben Goldacre, Liam Hart, George Hickman, Peter Inglesby, Steven Maude, Jessica Morley, Amir Mehrkar, Thomas O’Dwyer, Rebecca M Smith, Pete Stokes, Tom Ward, Jon Massey, Milan Wiedemann, Christopher Bates, Jonathan Cockburn, Sam Harper, Frank Hester, John Parry.

## Contributions

Conceptualisation: ALS, RH, HC, BMK; Data curation: ALS, SCJB, BG, ID, RS; Formal analysis: ALS, MW; Funding acquisition: BG; Investigation: ALS; Methodology: ALS, BMK, RH, HC, VS, CW, JQ, MJ; Project administration: BMK, BG, AM; Resources: RS, SCJB, BG, ID; Software: ALS, MW, SCJB, ID, RS; Supervision: BMK, BG, AS; Validation: ALS, MW; Visualisation: ALS; Writing - original draft: ALS; Writing - review & editing: All authors. ALS, JM, PI directly accessed and verified the underlying data reported in the manuscript. All authors gave final approval of the version to be published and agree to be accountable for all aspects of the work. All authors confirm that they had full access to all the data in the study and accept responsibility to submit for publication

## Software and reproducibility

Data management was performed using Python 3.8, with analysis carried out using R 4.0. Code for data management and analysis, as well as codelists, are archived online https://github.com/opensafely/waiting-list. All iterations of the pre-specified study protocol are archived with version control.

## Patient and public involvement and engagement

OpenSAFELY has involved patients and the public in various ways: we developed a public website that provides a detailed description of the platform in language suitable for a lay audience (https://opensafely.org); we have participated in two citizen juries exploring public trust in OpenSAFELY; we have co-developed an explainer video (https://www.opensafely.org/about/); we have patient representation who are experts by experience on our OpenSAFELY Oversight Board; we have partnered with Understanding Patient Data to produce lay explainers on the importance of large datasets for research; we have presented at various online public engagement events to key communities (e.g., Healthcare Excellence Through Technology; Faculty of Clinical Informatics annual conference; NHS Assembly; HDRUK symposium); and more. To ensure the patient voice is represented, we are working closely to decide on language choices with appropriate medical research charities (e.g., Association of Medical Research Charities). We will share information and interpretation of our findings through press releases, social media channels, and plain language summaries.

## Funding

The OpenSAFELY platform is principally funded by grants from:

- NHS England [2023-2025];
- The Wellcome Trust (222097/Z/20/Z) [2020-2024];
- MRC (MR/V015737/1) [2020-2021].

Additional contributions to OpenSAFELY have been funded by grants from:

- MRC via the National Core Study programme, Longitudinal Health and Wellbeing strand (MC_PC_20030, MC_PC_20059) [2020-2022] and the Data and Connectivity strand (MC_PC_20058) [2021-2022];
- NIHR and MRC via the CONVALESCENCE programme (COV-LT-0009, MC_PC_20051) [2021-2024];
- NHS England via the Primary Care Medicines Analytics Unit [2021-2024].

The views expressed are those of the authors and not necessarily those of the NIHR, NHS England, UK Health Security Agency (UKHSA), the Department of Health and Social Care, or other funders. Funders had no role in the study design, collection, analysis, and interpretation of data; in the writing of the report; and in the decision to submit the article for publication. MJ is funded by a National Institute for Health and Care Research (NIHR) Advanced Fellowship [NIHR301413]. The views expressed in this publication are those of the authors and not necessarily those of the NIHR, NHS or the UK Department of Health and Social Care.

## Information Governance and Ethical Approval

NHS England is the data controller of the NHS England OpenSAFELY COVID-19 Service; TPP is the data processor; all study authors using OpenSAFELY have the approval of NHS England^1^. This implementation of OpenSAFELY is hosted within the [EMIS environment which is] [TPP environment which is] [EMIS and TPP environments which are] accredited to the ISO 27001 information security standard and [is][are] NHS IG Toolkit compliant;**^2^** Patient data has been pseudonymised for analysis and linkage using industry standard cryptographic hashing techniques; all pseudonymised datasets transmitted for linkage onto OpenSAFELY are encrypted; access to the NHS England OpenSAFELY COVID-19 service is via a virtual private network (VPN) connection; the researchers hold contracts with NHS England and only access the platform to initiate database queries and statistical models; all database activity is logged; only aggregate statistical outputs leave the platform environment following best practice for anonymisation of results such as statistical disclosure control for low cell counts.**^3^** The service adheres to the obligations of the UK General Data Protection Regulation (UK GDPR) and the Data Protection Act 2018. The service previously operated under notices initially issued in February 2020 by the the Secretary of State under Regulation 3(4) of the Health Service (Control of Patient Information) Regulations 2002 (COPI Regulations), which required organisations to process confidential patient information for COVID-19 purposes; this set aside the requirement for patient consent.**^4^** As of 1 July 2023, the Secretary of State has requested that NHS England continue to operate the Service under the COVID-19 Directions 2020.**^5^** In some cases of data sharing, the common law duty of confidence is met using, for example, patient consent or support from the Health Research Authority Confidentiality Advisory Group.**^6^**

Taken together, these provide the legal bases to link patient datasets using the service. GP practices, which provide access to the primary care data, are required to share relevant health information to support the public health response to the pandemic, and have been informed of how the service operates.

1. The NHS England OpenSAFELY COVID-19 service - privacy notice. NHS Digital (Now NHS England). https://digital.nhs.uk/coronavirus/coronavirus-covid-19-response-information-governance-hub/the-nhs-england-opensafely-covid-19-service-privacy-notice (accessed 4 July 2023).
2. Data Security and Protection Toolkit - NHS Digital. NHS Digital (Now NHS England). https://digital.nhs.uk/data-and-information/looking-after-information/data-security-and-information-governance/data-security-and-protection-toolkit (accessed 4 July 2023).
3. ISB1523: Anonymisation Standard for Publishing Health and Social Care Data. NHS Digital (Now NHS England). https://digital.nhs.uk/data-and-information/information-standards/information-standards-and-data-collections-including-extractions/publications-and-notifications/standards-and-collections/isb1523-anonymisation-standard-for-publishing-health-and-social-care-data (accessed 4 July 2023).
4. 4. Coronavirus (COVID-19): notice under regulation 3(4) of the Health Service (Control of Patient Information) Regulations 2002 – general. 2022. https://www.gov.uk/government/publications/coronavirus-covid-19-notification-of-data-controllers-to-share-information/coronavirus-covid-19-notice-under-regulation-34-of-the-health-service-control-of-patient-information-regulations-2002-general--2 (accessed 5 July 2023).
5. 5. Secretary of State for Health and Social Care - UK Government. COVID-19 Public Health Directions 2020: notification to NHS Digital. https://digital.nhs.uk/about-nhs-digital/corporate-information-and-documents/directions-and-data-provision-notices/secretary-of-state-directions/covid-19-public-health-directions-2020 (accessed 4 July 2023).
6. Confidentiality Advisory Group. Health Research Authority. https://www.hra.nhs.uk/about-us/committees-and-services/confidentiality-advisory-group/ (accessed 4 July 2023).

## Supplementary Files

### Box 1: Glossary

**Referral to treatment (RTT) pathway (time on waiting list):** RTT pathways are consultant-led referrals for non-emergency services. It is initiated on the “clock start” date, and ends on the “clock stop” date.

**Non-RTT pathway:** Non-RTT pathways are non-consultant-led and planned care services such as physiotherapy, outpatient follow-ups, cancer surveillance, transplant follow ups, some diagnostics and chronic disease management.

**Consultant-led:** Treatment where a consultant has clinical responsibility. A consultant refers to a person contracted by a healthcare provider, has been appointed by a consultant appointment committee, and is a member of a Royal College or Faculty.

**Clock start (waiting list start date):** The clock starts when the patient is referred to a consultant-led service where the patient will be assessed and, if appropriate, receive treated; or an interface or referral management or assessment service, which may result in an onward referral to a consultant-led service.

**Clock stop (waiting list end date):** The clock can stop for treatment, which is the date when first definitive treatment starts; if the patient is admitted for treatment, the clock stop date is the date of admission. The clock doesn’t stop if:

- the patient is admitted for a diagnostic test / procedure only;
- the patient is admitted for pre-treatment;
- the patient is admitted for pre-op assessment only;
- the patient is admitted but doesn’t receive the intended procedure.

The clock can also stop for non-treatment; for instance, if the patient enters active monitoring, the patient declines treatment, a clinical decision is made not to treat, or the patient dies.

**First definitive treatment:** An intervention intended to manage a patient’s disease. What constitutes first definitive treatment is based on clinical judgment in consultation with the patient.

**Admitted pathway:** RTT pathways that end with admission for an inpatient or day case procedure. These are sometimes referred to as inpatient waiting times.

**Non-admitted pathway:** Waits that end for reasons other than an inpatient or day case admission to hospital for treatment. These are sometimes referred to as outpatient waiting times. They include patients whose RTT waiting time clock either stopped for outpatient treatment or for non-treatment.

Adapted from: *Recording and reporting referral to treatment (RTT) waiting times for consultant-led elective care* (https://www.england.nhs.uk/statistics/statistical-work-areas/rtt-waiting-times/rtt-guidance/)

---

**Supplementary Figure 1.**
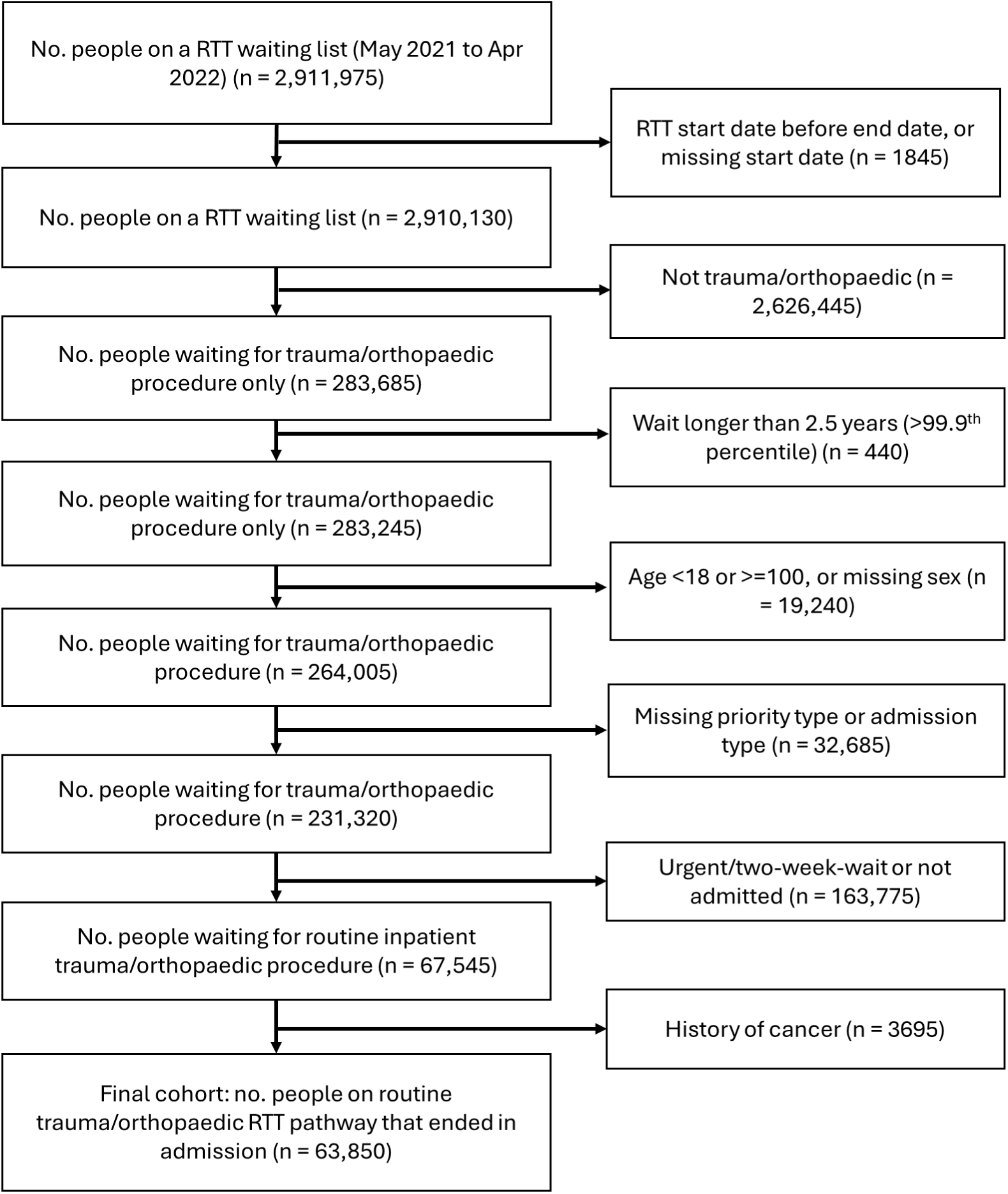
Flow chart with study population inclusion and exclusion criteria

**Supplementary Figure 2.**
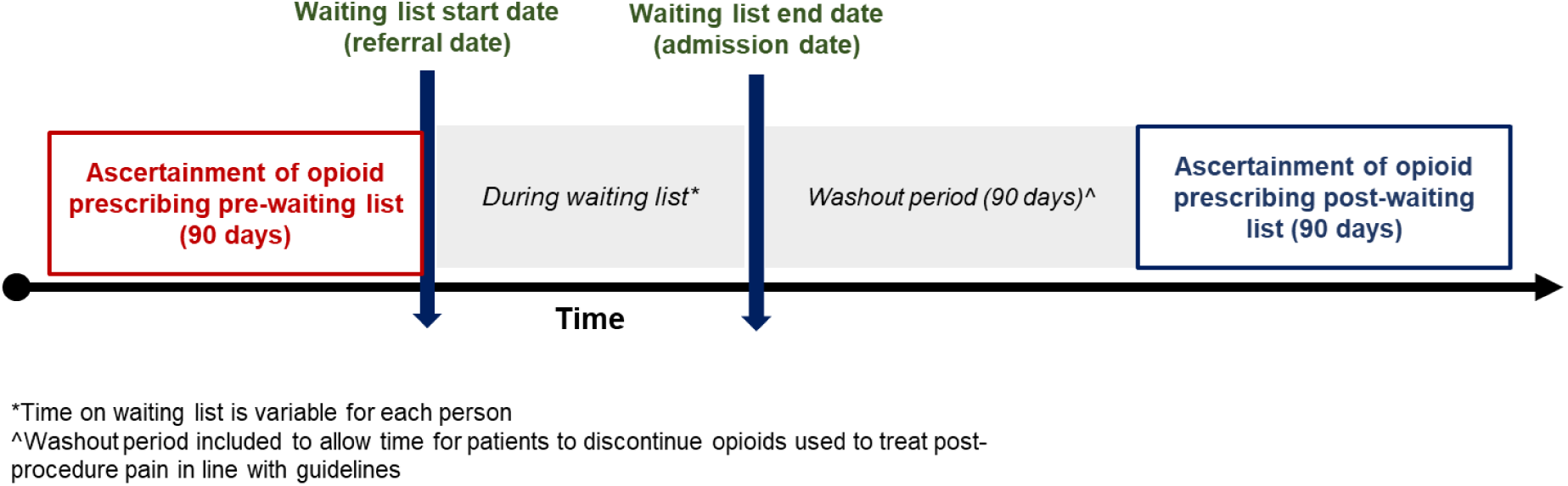
Visualisation of study design for comparison of opioid prescribing before and after waiting list

**Supplementary Table 1.** List of non-trauma orthopaedic procedure HRG codes.

| **Hip procedures** |  |
| --- | --- |
| HN12A | Very Major Hip Procedures for Non-Trauma with CC Score 10+ |
| HN12B | Very Major Hip Procedures for Non-Trauma with CC Score 8-9 |
| HN12C | Very Major Hip Procedures for Non-Trauma with CC Score 6-7 |
| HN12D | Very Major Hip Procedures for Non-Trauma with CC Score 4-5 |
| HN12E | Very Major Hip Procedures for Non-Trauma with CC Score 2-3 |
| HN12F | Very Major Hip Procedures for Non-Trauma with CC Score 0-1 |
| HN13A | Major Hip Procedures for Non-Trauma, 19 years and over, with CC Score 10+ |
| HN13B | Major Hip Procedures for Non-Trauma, 19 years and over, with CC Score 6-9 |
| HN13C | Major Hip Procedures for Non-Trauma, 19 years and over, with CC Score 4-5 |
| HN13D | Major Hip Procedures for Non-Trauma, 19 years and over, with CC Score 2-3 |
| HN13E | Major Hip Procedures for Non-Trauma, 19 years and over, with CC Score 1 |
| HN13F | Major Hip Procedures for Non-Trauma, 19 years and over, with CC Score 0 |
| HN13G | Major Hip Procedures for Non-Trauma, 18 years and under, with CC Score 1+ |
| HN13H | Major Hip Procedures for Non-Trauma, 18 years and under, with CC Score 0 |
| HN14A | Intermediate Hip Procedures for Non-Trauma, 19 years and over, with CC Score 6+ |
| HN14B | Intermediate Hip Procedures for Non-Trauma, 19 years and over, with CC Score 4-5 |
| HN14C | Intermediate Hip Procedures for Non-Trauma, 19 years and over, with CC Score 2-3 |
| HN14D | Intermediate Hip Procedures for Non-Trauma, 19 years and over, with CC Score 1 |
| HN14E | Intermediate Hip Procedures for Non-Trauma, 19 years and over, with CC Score 0 |
| HN14F | Intermediate Hip Procedures for Non-Trauma, between 6 and 18 years, with CC Score 1+ |
| HN14G | Intermediate Hip Procedures for Non-Trauma, between 6 and 18 years, with CC Score 0 |
| HN14H | Intermediate Hip Procedures for Non-Trauma, 5 years and under |
| HN15A | Minor Hip Procedures for Non-Trauma, 19 years and over |
| HN15B | Minor Hip Procedures for Non-Trauma, 18 years and under |
| HN16A | Minimal Hip Procedures, 19 years and over |
| HN16B | Minimal Hip Procedures, between 6 and 18 years |
| HN16C | Minimal Hip Procedures, 5 years and under |
| **Knee procedures** |  |
| HN22A | Very Major Knee Procedures for Non-Trauma with CC Score 8+ |
| HN22B | Very Major Knee Procedures for Non-Trauma with CC Score 6-7 |
| HN22C | Very Major Knee Procedures for Non-Trauma with CC Score 4-5 |
| HN22D | Very Major Knee Procedures for Non-Trauma with CC Score 2-3 |
| HN22E | Very Major Knee Procedures for Non-Trauma with CC Score 0-1 |
| HN23A | Major Knee Procedures for Non-Trauma, 19 years and over, with CC Score 4+ |
| HN23B | Major Knee Procedures for Non-Trauma, 19 years and over, with CC Score 2-3 |
| HN23C | Major Knee Procedures for Non-Trauma, 19 years and over, with CC Score 0-1 |
| HN23D | Major Knee Procedures for Non-Trauma, 18 years and under, with CC Score 1+ |
| HN23E | Major Knee Procedures for Non-Trauma, 18 years and under, with CC Score 0 |
| HN24A | Intermediate Knee Procedures for Non-Trauma, 19 years and over, with CC Score 4+ |
| HN24B | Intermediate Knee Procedures for Non-Trauma, 19 years and over, with CC Score 2-3 |
| HN24C | Intermediate Knee Procedures for Non-Trauma, 19 years and over, with CC Score 0-1 |
| HN24D | Intermediate Knee Procedures for Non-Trauma, between 6 and 18 years, with CC Score 1+ |
| HN24E | Intermediate Knee Procedures for Non-Trauma, between 6 and 18 years, with CC Score 0 |
| HN24F | Intermediate Knee Procedures for Non-Trauma, 5 years and under |
| HN25A | Minor Knee Procedures for Non-Trauma, 19 years and over |
| HN25B | Minor Knee Procedures for Non-Trauma, 18 years and under |
| HN26A | Minimal Knee Procedures, 19 years and over |
| HN26B | Minimal Knee Procedures, between 6 and 18 years |
| HN26C | Minimal Knee Procedures, 5 years and under |

**Supplementary Figure 3.**
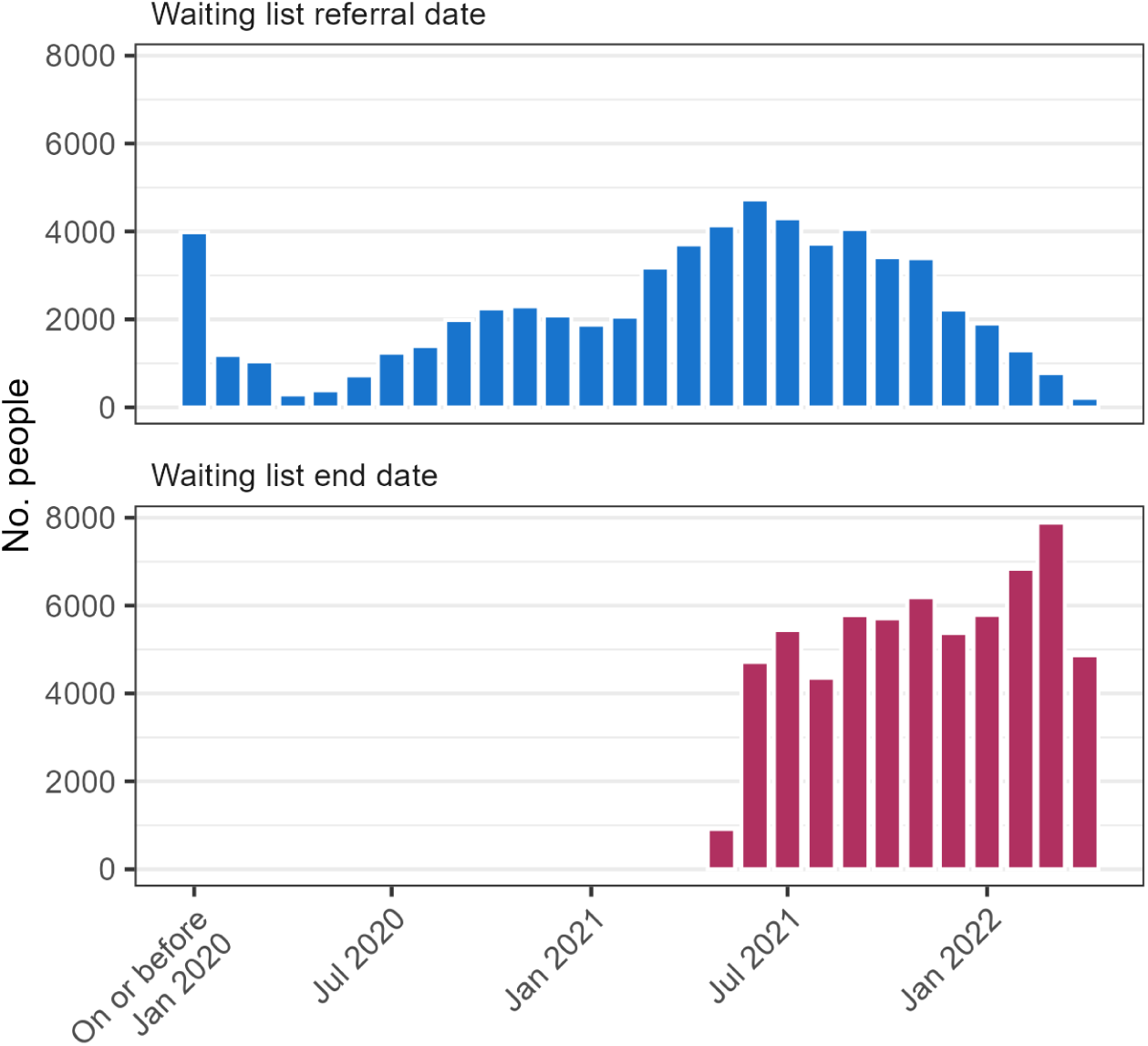
Distribution of waiting list start date (referral date) and end date (admission date) (n = 63,850)

**Supplementary Table 2.** Characteristics of people with recorded osteoarthritis diagnosis, and who received a hip or knee procedure for sensitivity analyses.

|  | Osteoarthritis<br>diagnosis | Hip procedure | Knee procedure |
| --- | --- | --- | --- |
|  | n (%) | n (%) | n (%) |
| **Total** | 24,255 (100.0) | 10,160 (100.0) | 14,975 (100.0) |
| **Time on waiting list** |  |  |  |
| <=18 weeks | 8370 (34.5) | 3220 (31.7) | 4755 (31.8) |
| 19-52 weeks | 10275 (42.4) | 4655 (45.8) | 6475 (43.2) |
| >52 weeks | 5610 (23.1) | 2285 (22.5) | 3745 (25.0) |
| **Age** |  |  |  |
| 18-39 | 430 (1.8) | 505 (5.0) | 2390 (16.0) |
| 40-49 | 1380 (5.7) | 650 (6.4) | 1515 (10.1) |
| 50-59 | 4855 (20) | 1720 (16.9) | 2890 (19.3) |
| 60-69 | 7305 (30.1) | 2780 (27.4) | 3605 (24.1) |
| 70-79 | 7620 (31.4) | 3285 (32.3) | 3565 (23.8) |
| 80+ | 2670 (11.0) | 1215 (12.0) | 1005 (6.7) |
| **Sex** |  |  |  |
| Female | 14355 (59.2) | 6105 (60.1) | 7375 (49.2) |
| Male | 9905 (40.8) | 4055 (39.9) | 7595 (50.7) |
| **IMD decile** |  |  |  |
| 1 (most deprived) | 1895 (7.8) | 655 (6.4) | 1330 (8.9) |
| 2 | 2205 (9.1) | 845 (8.3) | 1420 (9.5) |
| 3 | 2230 (9.2) | 865 (8.5) | 1365 (9.1) |
| 4 | 2360 (9.7) | 975 (9.6) | 1480 (9.9) |
| 5 | 2540 (10.5) | 1060 (10.4) | 1560 (10.4) |
| 6 | 2875 (11.9) | 1260 (12.4) | 1720 (11.5) |
| 7 | 2690 (11.1) | 1230 (12.1) | 1635 (10.9) |
| 8 | 2515 (10.4) | 1110 (10.9) | 1490 (9.9) |
| 9 | 2470 (10.2) | 1085 (10.7) | 1480 (9.9) |
| 10 (least deprived) | 2075 (8.6) | 900 (8.9) | 1210 (8.1) |
| Missing | 405 (1.7) | 175 (1.7) | 285 (1.9) |
| **Ethnicity** |  |  |  |
| White | 18675 (77) | 7790 (76.7) | 10825 (72.3) |
| Black | 525 (2.2) | 60 (0.6) | 500 (3.3) |
| South Asian | 160 (0.7) | 50 (0.5) | 145 (1.0) |
| Mixed | 80 (0.3) | 30 (0.3) | 85 (0.6) |
| Other | 75 (0.3) | 30 (0.3) | 85 (0.6) |
| Unknown | 4740 (19.5) | 2200 (21.7) | 3330 (22.2) |
| **Conditions recorded in primary care in past 5 years** |  |  |  |
| Anxiety (symptoms or diagnosis) | 2755 (11.4) | 925 (9.1) | 1595 (10.7) |
| Cardiac disease | 3575 (14.7) | 1325 (13) | 1570 (10.5) |
| Chronic kidney disease | 1200 (4.9) | 465 (4.6) | 500 (3.3) |
| Chronic respiratory disease | 1360 (5.6) | 540 (5.3) | 490 (3.3) |
| Depression (symptoms or diagnosis) | 3055 (12.6) | 1025 (10.1) | 1725 (11.5) |
| Diabetes | 2725 (11.2) | 890 (8.8) | 1330 (8.9) |
| Osteoarthritis | 24255 (100.0) | 6050 (59.5) | 7015 (46.8) |
| Rheumatoid arthritis | 555 (2.3) | 170 (1.7) | 290 (1.9) |
| **Prescribing in 3 months prior to waiting list** |  |  |  |
| Antidepressants |  |  |  |
| ≥1 prescription | 7055 (29.1) | 2515 (24.8) | 3290 (22) |
| ≥3 prescriptions | 4225 (17.4) | 1405 (13.8) | 1815 (12.1) |
| Amitriptyline/duloxetine |  |  |  |
| ≥1 prescription | 3455 (14.2) | 1250 (12.3) | 1335 (8.9) |
| ≥3 prescriptions | 1760 (7.3) | 605 (6.0) | 670 (4.5) |
| Gabapentinoids |  |  |  |
| ≥1 prescription | 995 (9.8) | 1125 (7.5) | 995 (9.8) |
| ≥3 prescriptions | 625 (6.2) | 740 (4.9) | 625 (6.2) |
| NSAIDs |  |  |  |
| ≥1 prescription | 4780 (19.7) | 2100 (20.7) | 2540 (17.0) |
| ≥3 prescriptions | 1705 (7.0) | 725 (7.1) | 730 (4.9) |
| **Died in 6 months following waiting list end date** | 140 (0.6) | 45 (0.4) | 35 (0.2) |

**Supplementary Table 3.** Opioid prescribing before and after time on waiting list among groups for sensitivity analyses.“Pre-waiting list” is 3 months prior to referral date, and “Post-waiting list” is months 4-6 after waiting list end date.

|  | ≥1 prescription |  | ≥3 more prescriptions |  |
| --- | --- | --- | --- | --- |
|  | Pre-waiting list | Post-waiting list* | Pre-waiting list | Post-waiting list* |
| **Osteoarthritis diagnosis** |  |  |  |  |
| Any opioid | 10,345 (42.7) | 8,065 (35) | 5,160 (21.3) | 4,395 (19.1) |
| Immediate-release opioid | 9,515 (39.2) | 7,270 (31.5) | 4,215 (17.4) | 3,550 (15.4) |
| Modified-release opioid | 1,750 (7.2) | 1,545 (6.7) | 1,210 (5.0) | 1,135 (4.9) |
| Weak opioid | 7,585 (31.3) | 5,565 (24.1) | 2,950 (12.2) | 2,475 (10.7) |
| Moderate opioid | 2,140 (8.8) | 1,660 (7.2) | 1,050 (4.3) | 915 (4.0) |
| Strong opioid | 1,945 (8) | 1,735 (7.5) | 1,265 (5.2) | 1,195 (5.2) |
| **Knee procedure** |  |  |  |  |
| Any opioid | 4,860 (32.5) | 3,890 (27.3) | 2,210 (14.8) | 1,980 (13.9) |
| Immediate-release opioid | 4,505 (30.1) | 3,560 (25) | 1,820 (12.2) | 1,660 (11.7) |
| Modified-release opioid | 695 (4.6) | 595 (4.2) | 470 (3.1) | 435 (3.1) |
| Weak opioid | 3,660 (24.4) | 2,785 (19.6) | 1,270 (8.5) | 1,185 (8.3) |
| Moderate opioid | 1,015 (6.8) | 820 (5.8) | 485 (3.2) | 420 (3.0) |
| Strong opioid | 750 (5.0) | 690 (4.8) | 485 (3.2) | 465 (3.3) |
| **Hip procedure** |  |  |  |  |
| Any opioid | 4,525 (44.5) | 2,925 (30.2) | 2,180 (21.5) | 1,530 (15.8) |
| Immediate-release opioid | 4,225 (41.6) | 2,630 (27.2) | 1,845 (18.2) | 1,220 (12.6) |
| Modified-release opioid | 665 (6.5) | 550 (5.7) | 425 (4.2) | 400 (4.1) |
| Weak opioid | 3,515 (34.6) | 2,090 (21.6) | 1,365 (13.4) | 880 (9.1) |
| Moderate opioid | 835 (8.2) | 530 (5.5) | 390 (3.8) | 295 (3.0) |
| Strong opioid | 765 (7.5) | 610 (6.3) | 470 (4.6) | 415 (4.3) |

\*People with complete follow-up

**Supplementary Figure 4.**
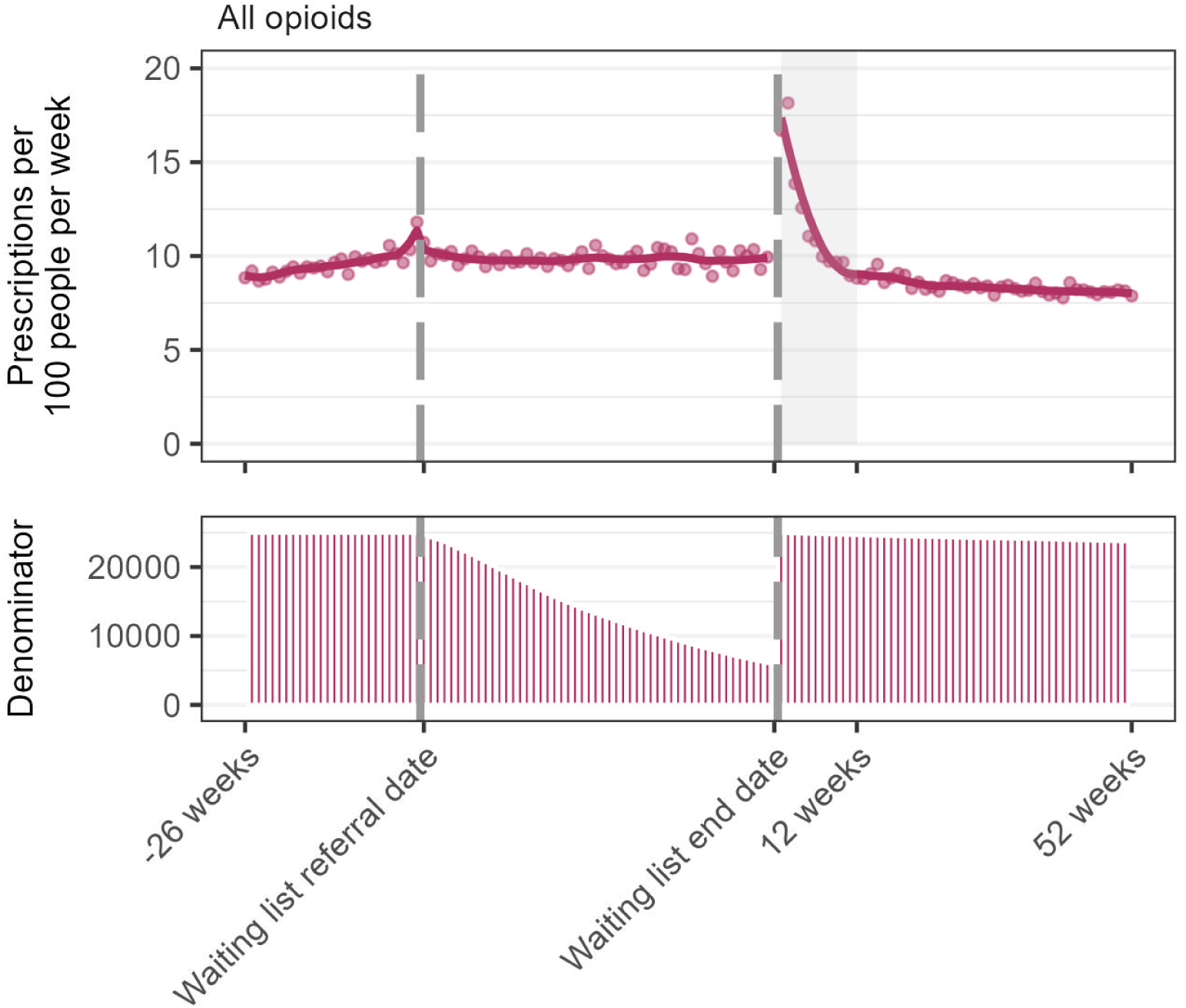
Number of opioid prescriptions per week in the 26 weeks prior to waiting list referral date, during the waiting list, and 52 weeks after waiting list end date among people with a **recorded diagnosis of osteoarthritis only**. Dots are observed values, solid lines are values predicted from loess regression model. Shaded area is 3 months post-admission. Legend: During the waiting list period, the denominator includes everyone who was still on the waiting list at the end of each week. During the waiting list period and post-waiting list periods, the denominator excludes people who died or who deregistered from their general practice.

**Supplementary Figure 5.**
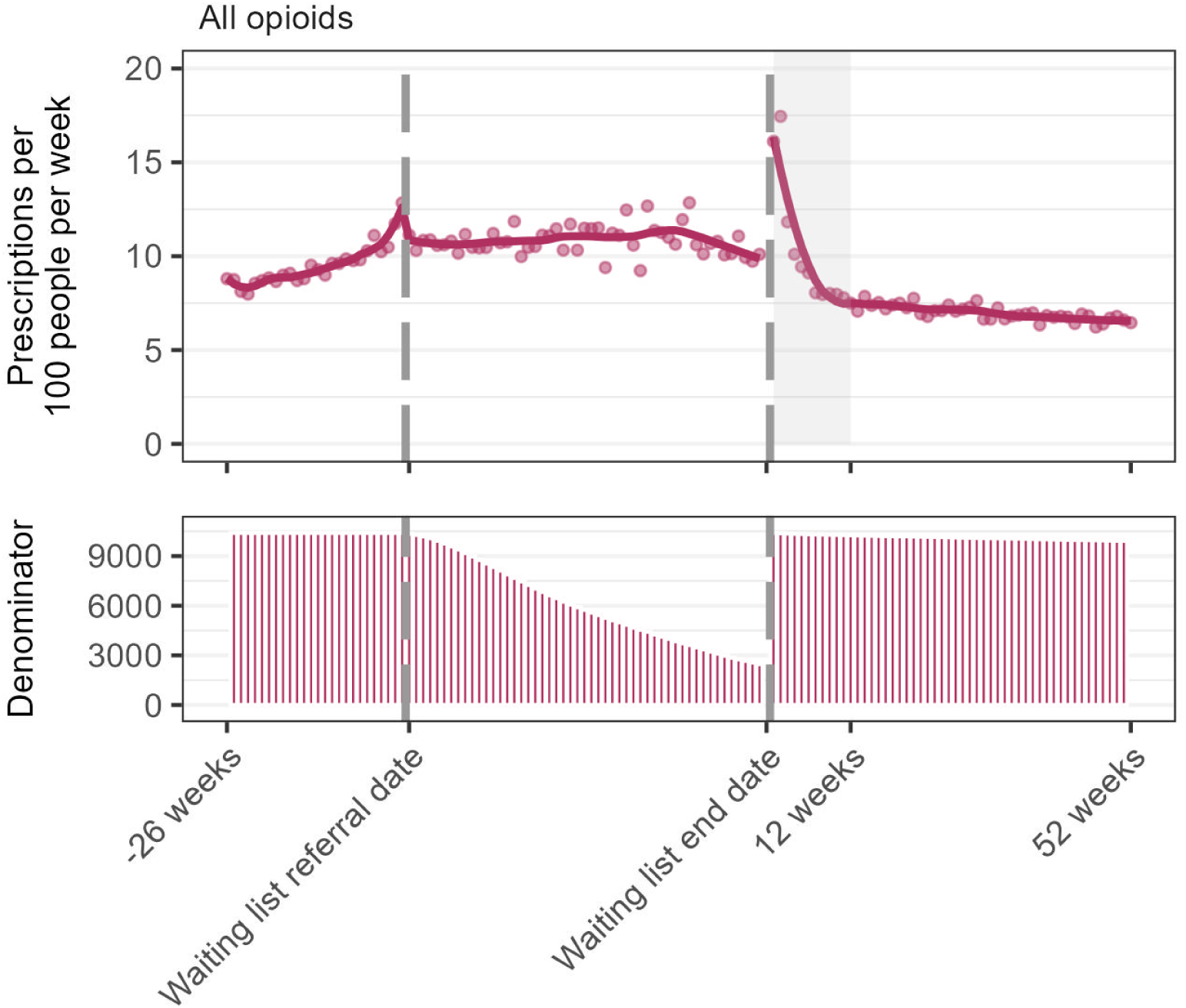
Number of opioid prescriptions per week in the 26 weeks prior to waiting list referral date, during the waiting list, and 52 weeks after waiting list end date among people who had a **hip procedure**. Dots are observed values, solid lines are values predicted from loess regression model. Shaded area is 3 months post-admission. Legend: During the waiting list period, the denominator includes everyone who was still on the waiting list at the end of each week. During the waiting list period and post-waiting list periods, the denominator excludes people who died or who deregistered from their general practice.

**Supplementary Figure 6.**
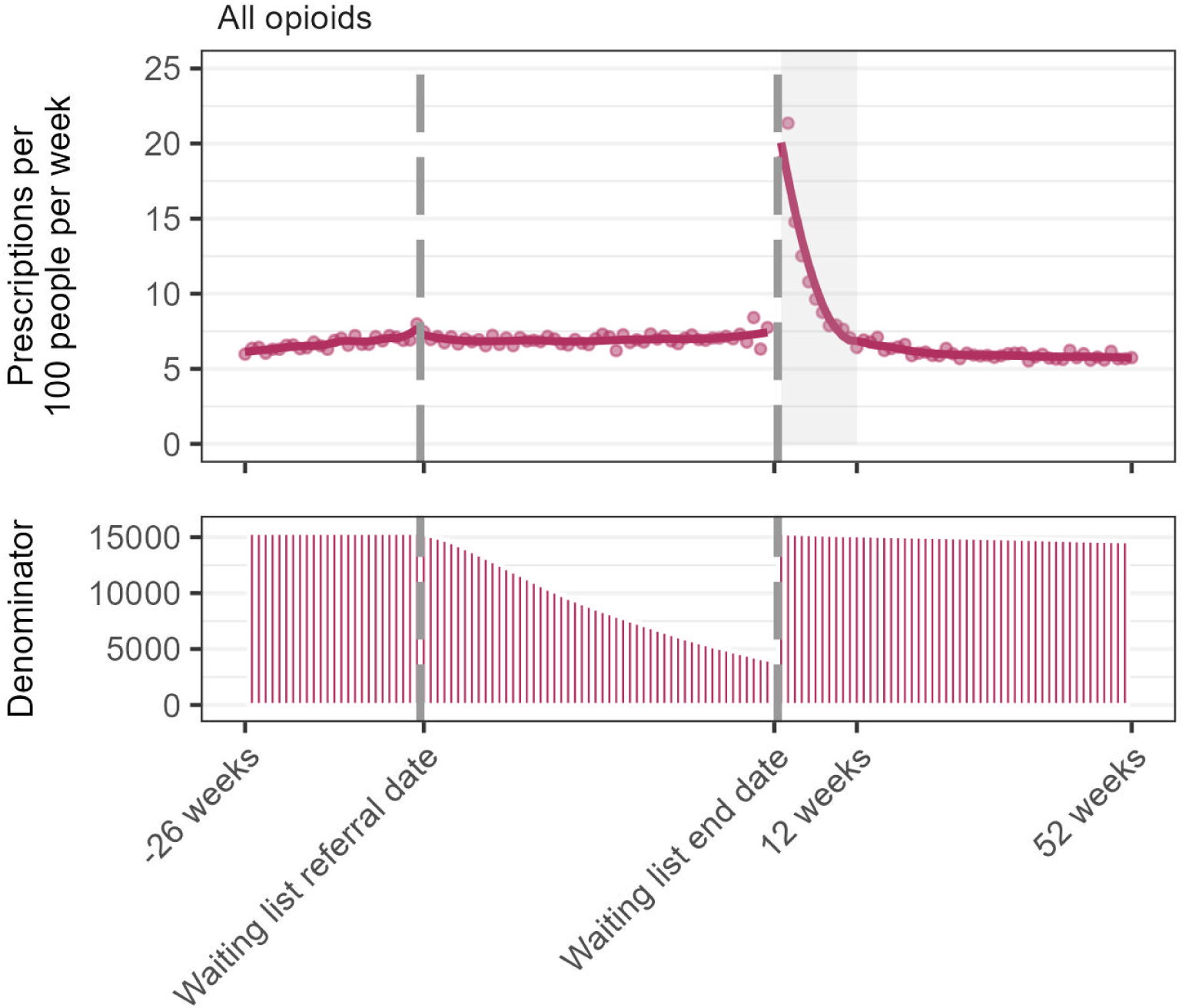
Number of opioid prescriptions per week in the 26 weeks prior to waiting list referral date, during the waiting list, and 52 weeks after waiting list end date among people who had a **knee procedure**. Dots are observed values, solid lines are values predicted from loess regression model. Shaded area is 3 months post-admission. Legend: During the waiting list period, the denominator includes everyone who was still on the waiting list at the end of each week. During the waiting list period and post-waiting list periods, the denominator excludes people who died or who deregistered from their general practice.

## References

1. British Medical Association. NHS backlog data analysis [Internet]. 2024 [cited 2024 Jan 31]. Available from: https://www.bma.org.uk/advice-and-support/nhs-delivery-and-workforce/pressures/nhs-backl og-data-analysis

2. The King’s Fund [Internet]. [cited 2023 Sep 28]. Waiting times for elective (non-urgent) treatment: referral to treatment (RTT). Available from: https://www.kingsfund.org.uk/projects/nhs-in-a-nutshell/waiting-times-non-urgent-treatment

3. Robertson R, Blythe N, Jefferies D. Tackling health inequalities on NHS waiting lists: learning from local case studies [Internet]. The King’s Fund; 2023 Nov [cited 2024 Jan 9]. Available from: https://www.kingsfund.org.uk/publications/health-inequalities-nhs-waiting-lists

4. Versus Arthritis. Versus Arthritis. 2023 [cited 2024 Apr 18]. The State of Musculoskeletal Health. Available from: https://versusarthritis.org/about-arthritis/data-and-statistics/the-state-of-musculoskeletal-health/

5. NHS Digital. NHS Digital. [cited 2024 Mar 5]. Hospital Admitted Patient Care Activity, 2021-22. Available from: https://digital.nhs.uk/data-and-information/publications/statistical/hospital-admitted-patient-care-activity/2021-22

6. Blom AW, Donovan RL, Beswick AD, Whitehouse MR, Kunutsor SK. Common elective orthopaedic procedures and their clinical effectiveness: umbrella review of level 1 evidence. BMJ. 2021 Jul 8;374:n1511.

7. Matharu G, Culliford D, Blom A, Judge A. Projections for primary hip and knee replacement surgery up to the year 2060: an analysis based on data from The National Joint Registry for England, Wales, Northern Ireland and the Isle of Man. Ann R Coll Surg Engl. 2022 Jun;104(6):443–8.

8. Farrow L, Gardner WT, Tang CC, Low R, Forget P, Ashcroft GP. Impact of COVID-19 on opioid use in those awaiting hip and knee arthroplasty: a retrospective cohort study. BMJ Qual Saf. 2021 Sep 14;bmjqs-2021-013450.

9. Rajamäki TJ, Puolakka PA, Hietaharju A, Moilanen T, Jämsen E. Use of prescription analgesic drugs before and after hip or knee replacement in patients with osteoarthritis. BMC Musculoskelet Disord. 2019 Sep 14;20(1):427.

10. Blågestad T, Nordhus IH, Grønli J, Engesæter LB, Ruths S, Ranhoff AH, et al. Prescription trajectories and effect of total hip arthroplasty on the use of analgesics, hypnotics, antidepressants, and anxiolytics: results from a population of total hip arthroplasty patients. PAIN. 2016 Mar;157(3):643.

11. Lee B, Yang KC, Kaminski P, Peng S, Odabas M, Gupta S, et al. Substitution of Nonpharmacologic Therapy With Opioid Prescribing for Pain During the COVID-19 Pandemic. JAMA Netw Open. 2021 Dec 10;4(12):e2138453.

12. Jani M, Birlie Yimer B, Sheppard T, Lunt M, Dixon WG. Time trends and prescribing patterns of opioid drugs in UK primary care patients with non-cancer pain: A retrospective cohort study. PLoS Med. 2020 Oct;17(10):e1003270.

13. Quinlan J, Levy N, Lobo DN, Macintyre PE. Preoperative opioid use: a modifiable risk factor for poor postoperative outcomes. Br J Anaesth. 2021 Sep 1;127(3):327–31.

14. NHS England. Referral to Treatment (RTT) Waiting Times [Internet]. 2024 [cited 2024 Mar 5]. Available from: https://www.england.nhs.uk/statistics/statistical-work-areas/rtt-waiting-times/

15. NHS England. Consultant-led Referral to Treatment Waiting Times Rules and Guidance [Internet]. 2021 [cited 2023 Sep 27]. Available from: https://www.england.nhs.uk/statistics/statistical-work-areas/rtt-waiting-times/rtt-guidance/

16. National Institute for Health and Care Excellence. Osteoarthritis in over 16s: diagnosis and management [Internet]. NICE; 2022 [cited 2024 Mar 1]. Available from: https://www.nice.org.uk/guidance/ng226

17. Yu D, Hellberg C, Appleyard T, Dell’Isola A, Thomas GER, Turkiewicz A, et al. Opioid use prior to total knee replacement: comparative analysis of trends in England and Sweden. Osteoarthritis Cartilage. 2022 Jun 1;30(6):815–22.

18. Royal College of Surgeons of England. Orthopaedic Surgery [Internet]. [cited 2024 Sep 11]. Available from: https://www.rcseng.ac.uk/careers-in-surgery/trainees/foundation-and-core-trainees/surgical-specialties/orthopedic-surgery/

19. Royal College of Surgeons of England. Major Trauma Surgery [Internet]. [cited 2024 Sep 11]. Available from: https://www.rcseng.ac.uk/standards-and-research/standards-and-guidance/service-standards/major-trauma-surgery/

20. NHS. NHS Standards Directory. [cited 2025 Apr 25]. National Waiting List Weekly Minimum Data Set. Available from: https://standards.nhs.uk/published-standards/national-waiting-list-weekly-minimum-data-set

21. Nab L, Schaffer A, Hulme W, DeVito NJ, Dillingham I, Wiedemann M, et al. OpenSAFELY: a platform for analysing electronic health records designed for reproducible research. 2024 Feb 13 [cited 2024 Feb 13]; Available from: https://osf.io/hj2sg

22. NHS England. Waiting List Minimum Data Set (WLMDS) Information [Internet]. [cited 2024 Jul 31]. Available from: https://www.england.nhs.uk/statistics/statistical-work-areas/rtt-waiting-times/wlmds/

23. Sullivan MD, Edlund MJ, Zhang L, Unützer J, Wells KB. Association Between Mental Health Disorders, Problem Drug Use, and Regular Prescription Opioid Use. Arch Intern Med. 2006 Oct 23;166(19):2087–93.

24. National Institute for Clinical Excellence. Treatment summaries. Analgesics. [Internet]. [cited 2024 Mar 6]. Available from: https://bnf.nice.org.uk/treatment-summaries/analgesics/

25. Faculty of Pain Medicine of the Royal College of Anaesthetists. Surgery and Opioids: Best Practice Guidelines 2021 [Internet]. [cited 2024 Mar 6]. Available from: https://fpm.ac.uk/surgery-and-opioids-best-practice-guidelines-2021

26. Levy N, Mills P. Controlled-release opioids cause harm and should be avoided in management of postoperative pain in opioid naïve patients. Br J Anaesth. 2019 Jun 1;122(6):e86–90.

27. National Institute for Clinical Excellence. Neuropathic pain in adults: pharmacological management in non-specialist settings [Internet]. NICE; 2013 [cited 2024 Mar 6]. Available from: https://www.nice.org.uk/guidance/cg173/chapter/Recommendations

28. Huang YT, Jenkins DA, Peek N, Dixon WG, Jani M. High frequency of long-term opioid use among patients with rheumatic and musculoskeletal diseases initiating opioids for the first time. Ann Rheum Dis. 2023 Aug;82(8):1116–7.

29. Parkin E. NHS maximum waiting time standards. 2024 Jul 31 [cited 2024 Jul 31]; Available from: https://commonslibrary.parliament.uk/research-briefings/cbp-8846/

30. The Lancet Rheumatology. Too long to wait: the impact of COVID-19 on elective surgery. Lancet Rheumatol. 2021 Feb;3(2):e83.

31. Schaffer AL, Andrews CD, Brown AD, Croker R, Hulme WJ, Nab L, et al. Changes in opioid prescribing during the COVID-19 pandemic in England: an interrupted time-series analysis in the OpenSAFELY-TTP cohort. Lancet Public Health. 2024 Jul;9(7):e432–42.

32. Huang YT, Jenkins DA, Yimer BB, Benitez-Aurioles J, Peek N, Lunt M, et al. Trends for opioid prescribing and the impact of the COVID-19 pandemic in patients with rheumatic and musculoskeletal diseases between 2006 and 2021. Rheumatol Oxf Engl. 2024 Apr 2;63(4):1093–103.

33. Lawal OD, Gold J, Murthy A, Ruchi R, Bavry E, Hume AL, et al. Rate and Risk Factors Associated With Prolonged Opioid Use After Surgery: A Systematic Review and Meta-analysis. JAMA Netw Open. 2020 Jun 25;3(6):e207367.

34. de Oliveira Costa J, Bruno C, Baranwal N, Gisev N, Dobbins TA, Degenhardt L, et al. Variations in Long-term Opioid Therapy Definitions: A Systematic Review of Observational Studies Using Routinely Collected Data (2000-2019). Br J Clin Pharmacol. 2021 Oct;87(10):3706–20.

35. Ashaye T, Hounsome N, Carnes D, Taylor SJC, Homer K, Eldridge S, et al. Opioid prescribing for chronic musculoskeletal pain in UK primary care: results from a cohort analysis of the COPERS trial. BMJ Open. 2018 Jun 1;8(6):e019491.

36. Andrews C, Schultze A, Curtis H, Hulme W, Tazare J, Evans S, et al. OpenSAFELY: Representativeness of electronic health record platform OpenSAFELY-TPP data compared to the population of England. Wellcome Open Res. 2022;7:191.

37. Hagger-Johnson G, Harron K, Fleming T, Gilbert R, Goldstein H, Landy R, et al. Data linkage errors in hospital administrative data when applying a pseudonymisation algorithm to paediatric intensive care records. BMJ Open. 2015 Aug 21;5(8):e008118.

38. Nirupa Dattani PDN and AMCUL (School of HS. Office for National Statistics. 2012 [cited 2024 Jul 31]. Linking maternity data for England 2007: methods and data quality. Available from: https://webarchive.nationalarchives.gov.uk/ukgwa/20160108045723mp_/http://www.ons.gov.uk/ons/rel/hsq/health-statistics-quarterly/no--53--spring-2012/linkage-of-maternity-hospital-episode-statistics-data.html

39. Coathup V, Macfarlane A, Quigley M. Linkage of maternity hospital episode statistics birth records to birth registration and notification records for births in England 2005-2006: quality assurance of linkage. BMJ Open. 2020 Oct 26;10(10):e037885.

40. Coronavirus. Coronavirus » Delivery plan for tackling the COVID-19 backlog of elective care [Internet]. 2022 [cited 2025 Apr 2]. Available from: https://www.england.nhs.uk/coronavirus/publication/delivery-plan-for-tackling-the-covid-19-backlog-of-elective-care/

41. PM sets out plan to end waiting list backlogs through millions more appointments - GOV.UK [Internet]. [cited 2025 Apr 2]. Available from: https://www.gov.uk/government/news/pm-sets-out-plan-to-end-waiting-list-backlogs-through-millions-more-appointments

42. NHS England. Optimising personalised care for adults prescribed medicines associated with dependence or withdrawal symptoms: Framework for action for integrated care boards (ICBs) and primary care [Internet]. 2023 [cited 2024 Feb 15]. Available from: https://www.england.nhs.uk/long-read/optimising-personalised-care-for-adults-prescribed-medicines-associated-with-dependence-or-withdrawal-symptoms/

43. James C, Denholm R, Wood R. The cost of keeping patients waiting: retrospective treatment-control study of additional healthcare utilisation for UK patients awaiting elective treatment. BMC Health Serv Res. 2024 Apr 30;24(1):556.

44. Federico CA, Wang T, Doussau A, Mogil JS, Fergusson D, Kimmelman J. Assessment of Pregabalin Postapproval Trials and the Suggestion of Efficacy for New Indications: A Systematic Review. JAMA Intern Med. 2019 Jan 1;179(1):90–7.

45. Gomes T, Juurlink DN, Antoniou T, Mamdani MM, Paterson JM, Brink W van den. Gabapentin, opioids, and the risk of opioid-related death: A population-based nested case–control study. PLOS Med. 2017 Oct 3;14(10):e1002396.

46. Cooper GM, Bayram JM, Clement ND. The functional and psychological impact of delayed hip and knee arthroplasty: a systematic review and meta-analysis of 89,996 patients. Sci Rep. 2024 Apr 5;14(1):8032.

47. Cisternas AF, Ramachandran R, Yaksh TL, Nahama A. Unintended consequences of COVID-19 safety measures on patients with chronic knee pain forced to defer joint replacement surgery. PAIN Rep. 2020 Dec;5(6):e855.

